# Quantifying Social Determinants of Health for Disease Prediction: A Multi-Level Approach Using Healthy People 2030 and All of Us Data

**DOI:** 10.1101/2025.09.02.25334565

**Authors:** Micah R. Hysong, Alisa K. Manning, Michael Green, Iain R. Konigsberg, Luciana B. Vargas, Jayati Sharma, Leslie Lange, Megan K. Shuey, LaShaunta M. Glover, Genevieve L. Wojcik, Sandra Lee, Laura M. Raffield, Sara J. Cromer, Polygenic Risk Methods Development (PRIMED) Consortium

## Abstract

Despite the growing recognition that social determinants of health (SDoH) play a prominent role in shaping health outcomes, inconsistent measures across health systems and research studies - and the absence of best practices for harmonizing, transforming, or combining variables - limits our ability to incorporate them into disease models. To address this gap, we applied the Healthy People 2030 (HP2030) framework to define, quantify, and incorporate SDoH into composite scores for disease prediction modeling, using individual-level surveys and area-level socioeconomic status (SES) measures from the American Community Survey in participants from the All of Us Research Program. We compared individual and area-level metrics of different complexity and composition and assessed associations between these SDoH metrics and nine chronic conditions, including asthma, diabetes, and prostate cancer. We further compared their predictive utility to that of commonly used metrics such as SES, area-level measures (such as a deprivation index), and self-identified race and ethnicity (SIRE). We find that diseases have distinct “social architectures,” with variation in the predictive strength of SDoH and the relative contributions of individual-versus area-level factors, prompting the development of disease-specific polysocial risk scores (PsRS). Many PsRS showed improved performance when both individual- and area-level data were included, with combined models often matching or outperforming models using SIRE alone. Lastly, we performed a Phenome-Wide Association Study, suggesting that the inclusion of SDoH in disease modeling could improve the prediction of ∼70% of examined phenotypes. Our findings highlight the value of incorporating SDoH into disease prediction models and position these measures as a more interpretable, actionable alternative to race and ethnicity.

## Introduction

A growing body of evidence highlights the substantial role of social determinants of health (SDoH) in shaping disease risk and progression. For example, neighborhood deprivation, literacy, and asthma education are associated with severity of asthma symptoms, health care utilization, hospitalization, and emergency department visits ^1–4^. Economic stability, education, health literacy, housing instability, loneliness, discrimination, and neighborhood socioeconomic status and violence have all been implicated in cardiovascular disease ^5,6^. Moreover, cardiovascular, cancer, and overall mortality in cancer survivors is influenced by psychological distress, economic stability, neighborhood and physical environment, social cohesion, and food insecurity ^7^. Associations such as these have prompted interest in incorporating SDoH as explanatory variables into disease prediction models ^8^, potentially in combination with other measures such as genetic variants and laboratory biomarkers.

Healthy People 2030 (HP2030) is a United States (US) initiative focused on improving health and well-being that defines social determinants of health (SDoH) as “the conditions in the environment where people are born, learn, work, play, worship, and age.” HP2030 organizes these determinants into five domains: economic stability (ES), education access and quality (education), social and community context (SCC), neighborhood and built environment (NBE), and health care access and quality (HCAU)^9^. While race and ethnicity have historically been used in predictive modeling to capture disparities, they often serve as imprecise proxies for SDoH and other societal and structural factors ^10^. Due to this increased recognition, some studies have begun directly incorporating socioeconomic status (SES) metrics such as income and education into disease prediction models; however, these may not be sufficient to fully capture an individual’s SDoH and assume no heterogeneity of effect of SDoH in different populations ^11^. As more comprehensive SDoH data become available, and their relationship with clinical outcomes across populations more understood, SDoH metrics may be able to replace race and ethnicity in predictive modeling to more directly capture the proximal causes of health inequities and to avoid misinterpretation of associations leading to racial essentialism ^12^. Moreover, SDoH may be more modifiable and preventable than downstream clinical factors, making them good targets for population-level interventions ^13^. However, large-scale integration of SDoH in disease prediction modeling and clinical research remains challenging due to disparate measures across healthcare systems and research studies, the absence of defined gold standard measures and best practices for transforming or combining variables, and limited understanding of how individual-level and area-level factors contribute to disease risk across populations and for different pathologies, resulting in limited reproducibility and generalizability of results ^14,15^.

To address this gap, our study applied the HP2030 framework to define, quantify, and incorporate numerous SDoH into predictive models of disease for nine chronic conditions - including asthma, prostate cancer, and diabetes - previously prioritized for clinical risk modeling ^16^. Using individual-level survey data from the All of Us Research Program, we constructed five domain-specific SDoH scores aligned with the HP2030 framework, and an overall composite SDoH score to provide a comprehensive measure of social risk from individual-level surveys. We then examined the correlation of these scores and evaluated their associations with nine chronic conditions, comparing their performance against often more readily available individual SES variables, area-level metrics, and self-identified race and ethnicity ^16^. Our approach explores the strengths, limitations, and interrelatedness of a variety of SDoH measures; allows for the investigation of disease-specific patterns of SDoH association; and highlights how considering both individual-level and area-level SDoH can enhance disease prediction across a diverse cohort. We further demonstrate that rich individual-level SDoH data can be used to reduce reliance on race and ethnicity in predictive modeling.

## Methods

### All of Us Research Program

The All of Us Research Program (AoU) - a National Institutes of Health (NIH)-funded initiative designed to increase the scale and diversity of biomedical research participants and to reduce health disparities - began enrolling participants in May 2018^17^. The program implements multiple strategies to ensure adequate representation of individuals historically underrepresented in biomedical research and collects a wide range of data, including demographic characteristics, individual- and area-level SDoH, electronic health records (EHRs), and genetic information. Version release 8 (V8) includes 633,547 individuals.

### SDoH instruments

As a part of its diverse data collection to advance precision health, AoU administers surveys to its participants. The Basics survey was among the first three surveys developed by All of Us and is administered at enrollment ^18^. It collects core demographic information, including self-reported race, ethnicity, income, and education. 633,532 participants completed the Basics survey in V8 (the Full Cohort), although with various degrees of item non-response (17.3% for race and ethnicity, 2.3% for educational attainment, 18.1% for income). AoU also employed a SDoH Task Force of subject matter experts, who developed a scientifically valid and reliable survey to collect self-reported data on key dimensions of SDoH ^19^. At the time of V8, 259,189 participants had completed at least some questions on the SDoH survey. There is non-response and non-random item missingness by educational attainment, racial and ethnic identity, and survey language ^19^. AoU also has a Health Care Access & Utilization (HCAU) survey, which 305,857 participants have completed.

AoU provides area-level SDoH measures at the three-digit zip code level through linkage with the 2017 American Community Survey (ACS). These include measures related to each HP2030 domain except social and community context (SCC), including economic stability (ES; median household income, percent receiving assisted income, percent living below the poverty limit), education access and quality (Education; percent of adults over 25 with a high school diploma), neighborhood and built environment (NBE; percent vacant housing), and health care access and quality (HCAU; percent without health insurance). Additionally, AoU includes the Nationwide Community Deprivation Index (NCDI), the first principal component from the six different ACS measures, which explains over 60% of the total variance in census-tract level measurements from the ACS ^20^. High school education and median household income were flipped to be in the direction of risk along with the rest of the measures.

### Cohorts

162,193 study participants completed the Basics survey and had adequate EHR completeness, defined as having at least three distinct clinical encounters over a span of three or more years. Among these, 125,295 had linked area-level SDoH data, education, income, and household size data (“SES Cohort”), and 54,313 completed the individual-level SDoH and HCAU surveys with at least a 60% response rate across all five Healthy People 2030 domains, including income data for the economic stability domain (the “Individual Cohort”; Supplementary Figure 1).

Some analyses incorporated self-identified race and ethnicity (SIRE), which was categorized into three groups: non-Hispanic Black (NHB), non-Hispanic White (NHW), and Hispanic (HS). These were the only groups with sufficient sample sizes across all disease outcomes to support stratified analyses; therefore, individuals who self-identified as Middle Eastern or North African, multiracial, Native Hawaiian or Other Pacific Islander, Asian, or who skipped or declined to answer the demographic question were excluded from these analyses. This filtering resulted in the creation of two additional sub-cohorts derived from the SES and Individual Cohorts - limited to individuals identifying as NHW, NHB, or HS - the "SES-SIRE Cohort" (n = 117,535) and the "Individual-SIRE Cohort" (n=51,265).

### SDoH Domain Development

SDoH survey scores were derived from items included in the Basics, SDoH, and HCAU surveys (https://www.researchallofus.org/data-tools/survey-explorer/), following the mapping strategy outlined by the AoU SDOH Task Force and hosted on AoU as a demonstration workspace (“Demo - Social Determinants of Health;” ^19^. This framework was applied to the SDoH survey, and a similar approach, with additional guidance from the PhenX Toolkit, was used to construct survey scales from the HCAU survey ^21^. We calculated Cronbach’s alpha to ensure internal consistency of each scale of related survey items within our cohort. “Is there a place that you USUALLY go to when you are sick or need advice about your health?” was excluded due to the low number of “No” responses (∼2%) in our Individual Cohort, likely related to cohort restriction to those with sufficient EHR depth. Further methodological details, including transformation of survey items and scale development information, are provided in Appendix I.

SDoH instrument scales were grouped into domains following the HP2030 framework and single latent domain scores were derived for each domain using Confirmatory Factor Analysis (CFA) with the lavaan package (missing = "fiml") (version 0.6-19) in R (version 4.5.0). Full Information Maximum Likelihood (FIML) was used to handle missing data. Additionally, CFA was used to derive an overall composite SDoH metric from the five SDoH domains. For the composite metric, social cohesion was cross-loaded between SCC and NBE as it comes from a social cohesion among neighbors survey. Employment-related items were excluded from the financial security domain, as they did not adequately capture this construct in our Individual Cohort (reduced model fit). A combination of theory (conceptually related items) and modification indices (residual variance >0.09) were used to determine whether or not to include error correlations among related items (Figure 1). Model fit was assessed using comparative fit index (CFI), Root mean square error of approximation (RMSEA), and standardized root mean square residual (SRMR) where CFI > 0.95, RMSEA < 0.06, and SRMR < 0.10 constitute good model fit ^22,23^.

**Figure 1:**
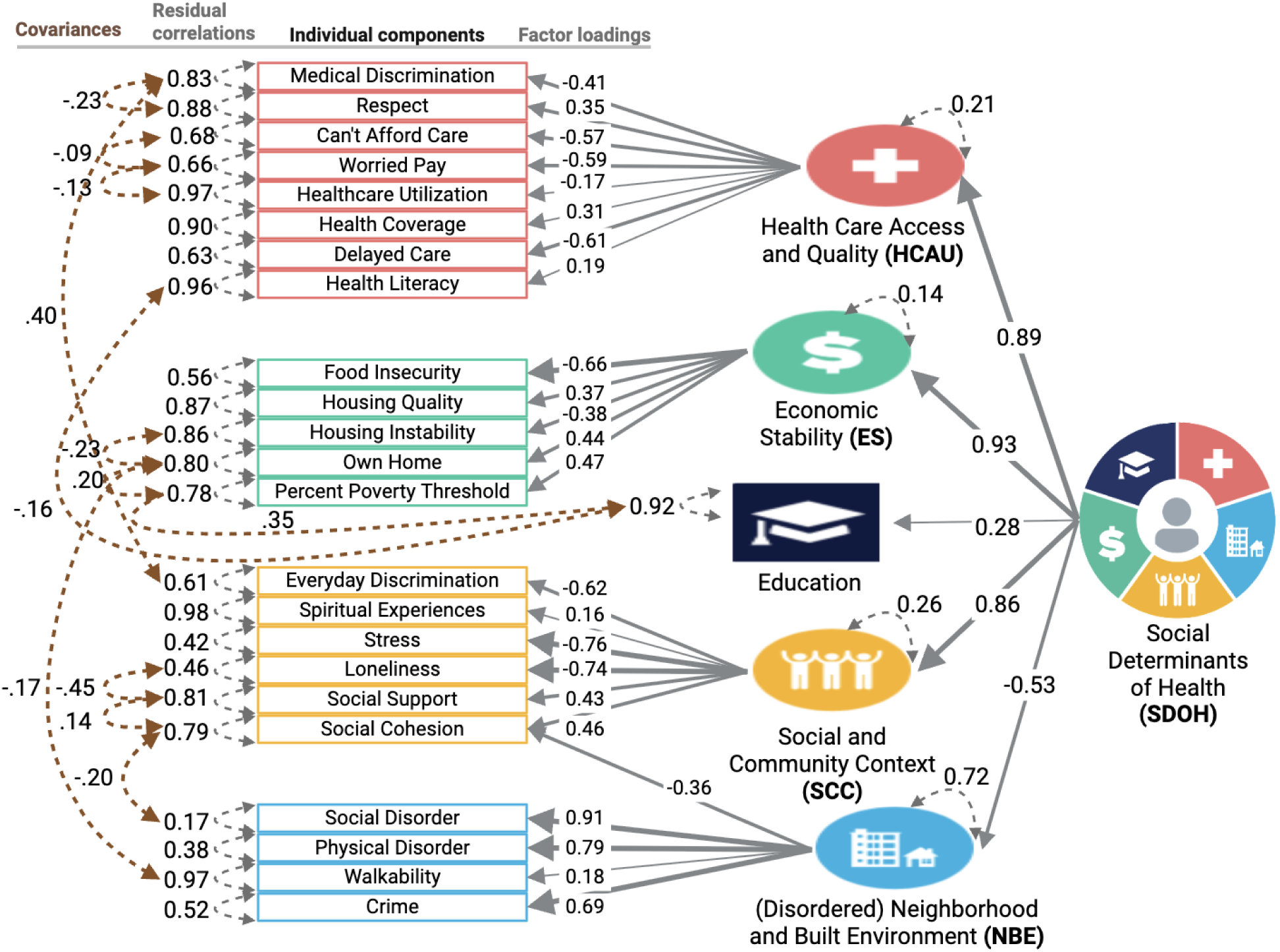
Social Determinants of Health Measurement Model(s) CFA models for four SDoH domains identified by HP 2030, alongside a higher-order model representing overall social advantage. Constructs are grouped and color-coded by domain. Grey single-headed arrows indicate standardized factor loadings, pointing left toward the observed variables, which serve as indicators of their respective latent constructs (HP2030 domains). Grey dashed double-headed arrows denote residual correlations, while brown arrows indicate covariances between item errors for closely related constructs.

### Disease algorithms

To evaluate how these latent SDoH constructs relate to health outcomes, we analyzed nine chronic conditions previously selected for their high prevalence, cost, and medical actionability ^16,24,25^. Disease definitions were adapted from validated EHR-based algorithms from the Electronic Medical Records and Genomics (eMERGE) network.

These algorithms are for asthma, atrial fibrillation (Afib), breast cancer, chronic kidney disease (CKD), coronary heart disease (CHD), hypercholesterolemia, prostate cancer, type 1 diabetes (T1D), and type 2 diabetes (T2D).

### Covariates

Sex at birth and gender (henceforth “Sex/Gender”) were self-reported and categorized as cisgender female, cisgender male, or a collapsed sexual or gender minority (SGM) categorization (aggregated due to limited sample size). Age was recorded at the last event in the EHR record. We approximated record depth by adding the number of unique visits in which the EHR record contained an observation or condition code. We then calculated visit frequency by dividing record depth over EHR length (max – min date in the record).

### Structural Equation Modeling (SEM)

Each disease outcome was modeled using Structural Equation Modeling (SEM) with a Weighted Least Squares Mean and Variance adjustment (WLSMV) using the *lavaan* package (version 0.6-19). To enable direct comparison of coefficients, all SDoH domain scores were min-max normalized to a 0–1 scale, with higher values indicating greater social adversity. Separate models were run for each individual-level and area-level SDoH domain with disease outcome, adjusting for age, age-squared, Sex/Gender, record depth, and visit frequency. Statistical significance was assessed using a Bonferroni-corrected threshold of P < 4.27 x 10^-4^ (0.05/117), accounting for 13 SDoH domains across 9 disease outcomes.

For breast cancer and prostate cancer models, we excluded participants identifying as cisgender males or cisgender females, respectively. Additionally, individuals identifying as SGM were excluded from CKD and T1D models due to insufficient case counts (<20).

### Polysocial Risk Score (PsRS) Construction

PsRS were constructed using elastic net models implemented with cv.glmnet (version 4.1.8) in R with α = 0.5. Each elastic net model was trained using a 70/30 training/testing split and evaluated using five-fold cross-validation. For each fold, the λ value minimizing lowest mean cross-validation error was extracted, and the mean and standard deviation across the five folds were calculated.

PsRS were constructed for the following sets of predictors in the Individual-SIRE Cohort (n=51,265):

1. **Domain PsRS**: individual-level SDoH domain composite scores (n=6)
2. **\*Component PsRS:** SDoH domain composite scores (n=6) and components of SDoH domain composite scores (n=23)
3. **Area PsRS:** area-level predictors (n=7)
4. **Combined PsRS:** SDoH domain composite scores (n=6), components of SDoH domain composite scores (n=23), and area-level predictors (n=7)

PsRS were constructed for the following sets of predictors in the SES-SIRE Cohort (n = 117,535):

1. **Area PsRS:** area-level predictors (n=7)
2. **Combined PsRS:** percent of poverty threshold, education, and area-level predictors (n=7)

*For the Component PsRS, the mice package in R was used to impute missing data with 10 imputations using predictive mean matching (ridge = 0.001). Two variables had no missingness, five had less than 1% missingness, six had less than 2%, six had less than 5%, and two had less than 10%. The variables with the highest rates of missingness were Delayed Care (11.48%) and Healthcare Utilization (17.96%). All variables were used in the imputation of others, as well as the covariates and SIRE (due to the correlation of SIRE with SDoH).

### Disease prediction model evaluation

Standard linear regression models were fitted, and model performance was evaluated using the area under the curve (AUC). Ninety-five percent confidence intervals for the AUC were calculated and reported. NHW was set as the reference group in relevant analyses since it is the largest sample. Statistical differences between models were assessed using roc.test from the pROC package (version 1.19.0.1) in R with DeLong’s method. A Bonferroni threshold of P < 1.54 x 10^-04^ (0.05/324) was used to adjust for the number of comparisons in the Individual-SIRE Cohort, P < 2.65 x 10^-04^ (0.05/189) in the SES-SIRE Cohort, and P < 1.85 x 10^-03^ (0.05/81) in the stratified analysis.

Nine sets of predictors were compared in the Individual-SIRE Cohort (n=51,265):

1. **Base model:** age (at last EHR entry), age^2^, Sex/Gender
2. **EHR model:** Base model + visit frequency + record depth
3. **SIRE model**: EHR model + NHB + HS
4. **SES model:** EHR model + percent of poverty threshold + education
5. **Domain PsRS model:** EHR model + Domain PsRS
6. **Component PsRS model:** EHR model + Component PsRS
7. **Area PsRS model:** EHR model + Area PsRS
8. **Combined PsRS model:** EHR model + Combined PsRS
9. **Combined PsRS+ model:** EHR model + Combined PsRS + NHB + HS

Seven sets of predictors were compared in the SES-SIRE Cohort (n = 117,535):

1. **Base model:** age, age^2^, Sex/Gender
2. **EHR model:** Base model + visit frequency + record depth
3. **SIRE model**: EHR model + NHB + HS
4. **SES model:** EHR model + percent of poverty threshold + education
5. **Area PsRS model:** EHR model + Area PsRS
6. **Combined PsRS model:** EHR model + Combined PsRS
7. **Combined PsRS+ model:** EHR model + Combined PsRS + NHB + HS

Additionally, Base, EHR, and Combined models were run in analyses stratified by SIRE (NHB, NHW, HS) in the SES-SIRE Cohort to assess intersectionality of SIRE and SDoH.

### Phenome-Wide Association Study (PheWAS)

PheWAS can be used to scan for associations with groups of International Classification of Diseases (ICD) codes (phecodes) to capture meaningful relationships between predictors and disease ^26^. We adapted the PheWAS pipeline hosted by AoU as a demonstration workspace (“Demo - PheWAS Smoking”); using phecode v1.2; setting the minimum number of cases to 100 to ensure sufficient sample size and statistical power; and using age at most recent phecode, Sex/Gender, record depth, and visit frequency as covariates ^27^. Disease status was defined by the presence of at least two instances of the same phecode; individuals with only one instance were considered neither a case nor a control and were excluded from analysis for that disease. Phecode groupings were restricted to circulatory system, endocrine/metabolic, genitourinary, neoplasms, and respiratory, groupings also represented in our nine chronic conditions from main models.

A targeted PheWAS was conducted in each cohort. In the Individual Cohort, associations were tested across the five domains, the SDoH composite score, and the area-level metrics for a total of 13 predictors. 513 unique diseases were tested, resulting in a Bonferroni-corrected threshold of 7.50 x 10^-6^ (0.05/6669). In the SES Cohort, associations were tested across percent of poverty threshold, income, and the area-level metrics for a total of nine predictors. 621 unique diseases were tested, resulting in a Bonferroni-corrected threshold of 8.85 x 10^-6^ (0.05/5589).

All modeling was performed in R (version 4.5.0) and analysis scripts are publicly available on the All of Us research platform as a community workspace: “Social Determinants of Health Measurement Models V8”.

## Results

### Participant Characteristics

Among 633,547 participants in AoU V8 (the Full Cohort), 162,193 had sufficient electronic health records (EHRs). Among these, 125,295 had linked area-level SDoH data, individual-level income, and individual-level education data (SES Cohort). 54,313 had sufficient individual-level SDoH (Individual Cohort; Supplementary Figure 1). Notably, the individual-level SDoH survey data is subject to sampling bias, with older, White, higher-income, more highly educated, privately or Medicare-insured participants, and participants born in the USA more likely to have completed the survey (Table 1). Moreover, the Individual Cohort exhibits more favorable area-level SDoH characteristics compared to the SES Cohorts, which in turn show more favorable characteristics than the Full Cohort (Supplementary Table 1 S1). Within the Individual Cohort, the mean age was 59.4 years old (yo), 61.2% identified as cisgender female, 1.8% as Asian, 7.7% as Black, 1.5% with multiple races, 0.4% as Middle Eastern, 81.3% as White, and 6.7% as Hispanic (HS). A substantial 66.2% held at least a four-year college degree, compared to 37.7% of the US population in 2022 ^28^. Additionally, 21% reported annual household incomes of over 150k, and almost all had health insurance, with far fewer being covered by Medicaid compared to the Full Cohort. Overall, the Individual Cohort experiences a favorable distribution of individual-level SDoH characteristics, with most having healthcare coverage and low neighborhood crime and disorder (Supplementary Figure 2).

**Table 1:**
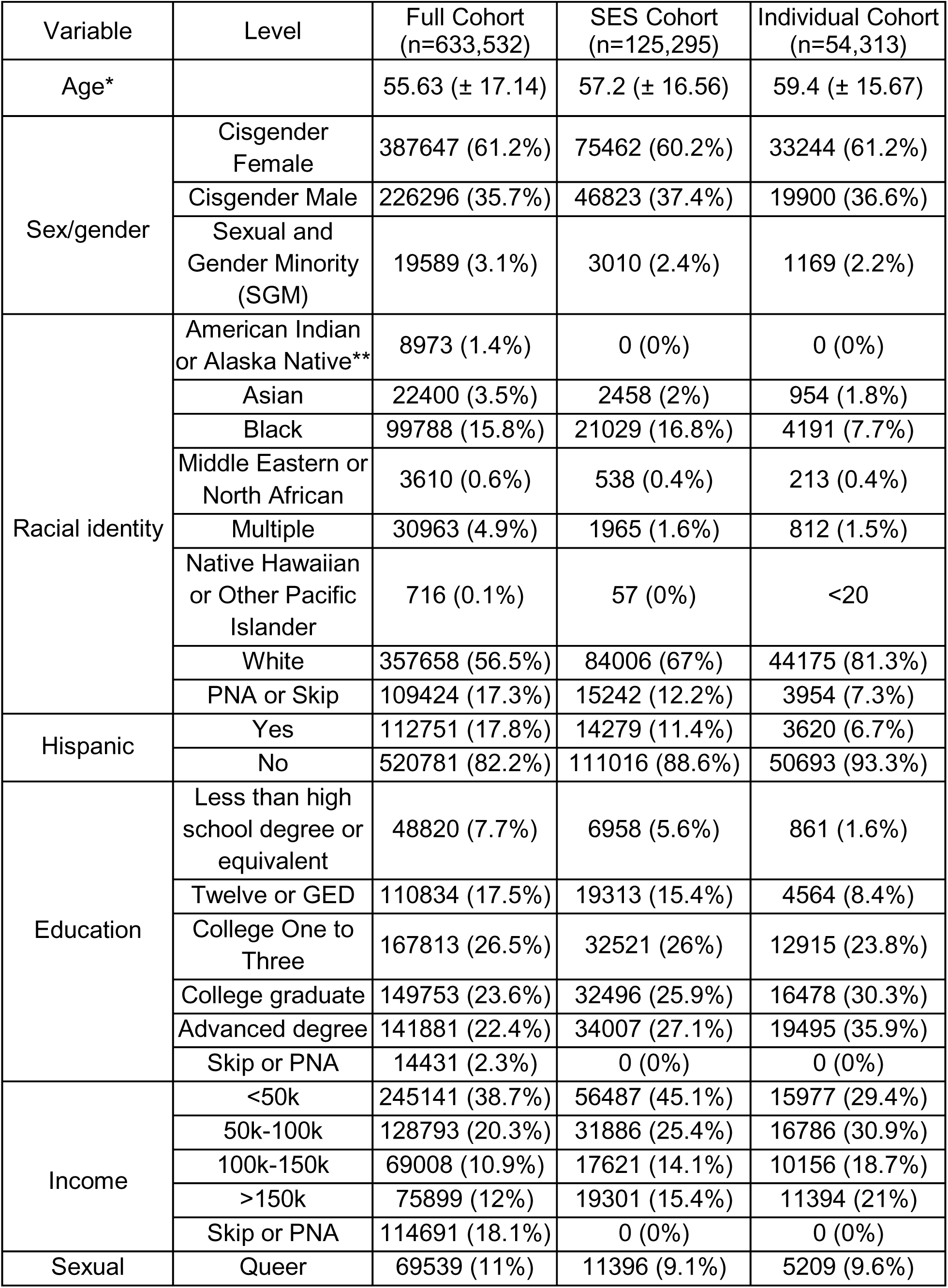
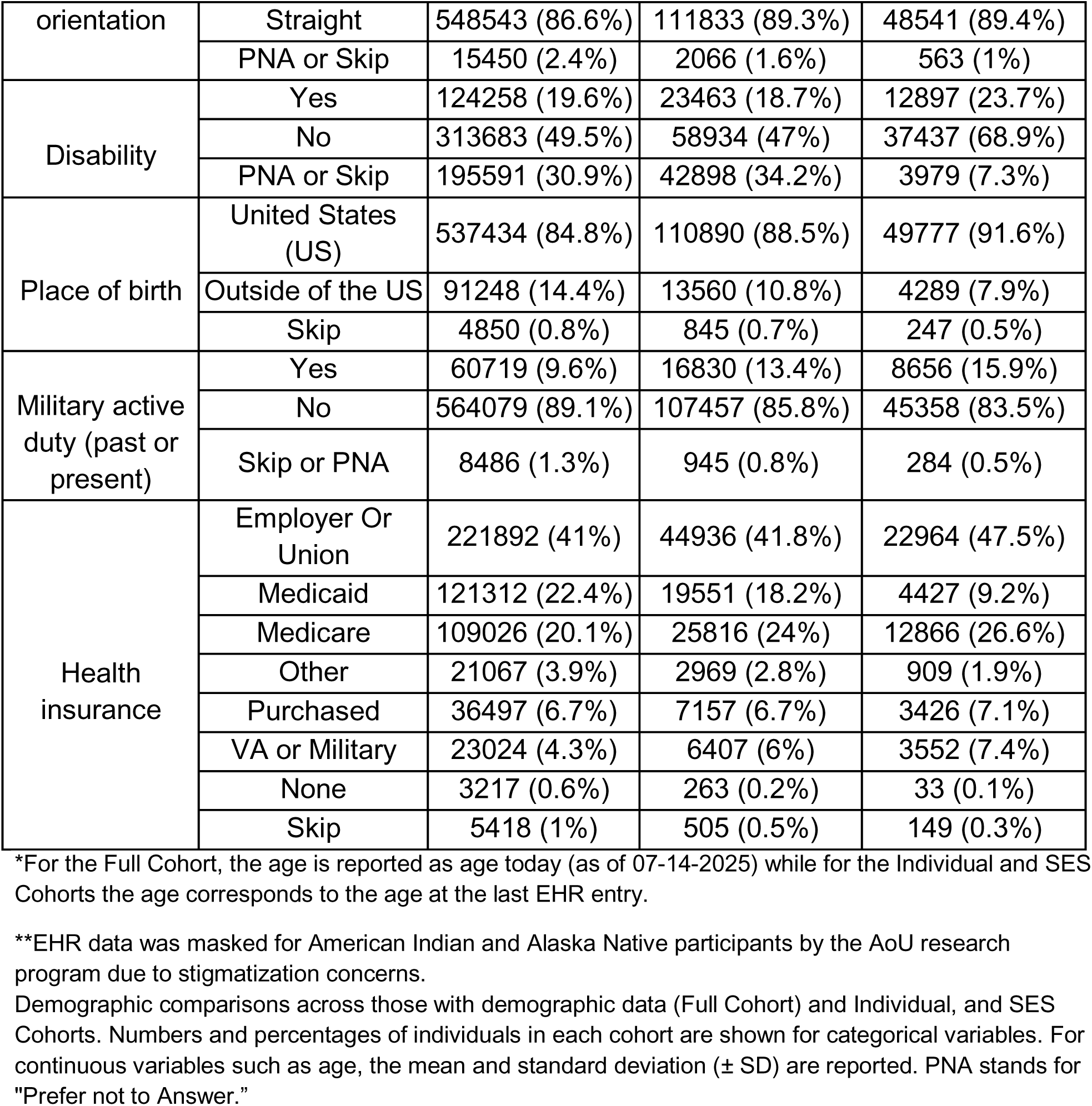
Cohort Demographic Comparisons.

Analysis of participant characteristics across SIRE groups reveals key patterns of demographic intersectionality. Participants identifying as non-Hispanic Black (NHB) and HS were consistently younger than other groups, while NHW participants were the oldest across all cohorts. Across all cohorts, NHB and HS participants had median ages of 57-58 yo and 51-52 yo, respectively, compared to 63-64 yo for NHW participants (Supplementary Table 1 S2). Additionally, NHW participants had the longest median record depth (107 encounters in the Full Cohort), while HS and NHB participants had shorter records, with median depths of 78 and 84 encounters, respectively. In the Individual Cohort, all groups except NHW exhibited evidence of selection bias favoring individuals with longer EHR records. While NHW participants saw only a modest increase in median record depth (from 107 to 109), the increases were more substantial for HS (from 78 to 91.5) and NHB (from 84 to 106.5) participants. Selection bias related to education and income were also more pronounced among NHB and HS groups. For example, the proportion of NHB and HS participants with an advanced degree increased from 5.1% to 11.5% and 5.7% to 11.6%, respectively, compared to a smaller increase among NHW participants—from 24.4% to 35.6%. Demographic information for case and control groups can be found in Supplementary Table 1 S3:4.

### Creation of SDoH Domain Scores

For individual-level survey data, a single latent score was derived for four out of five SDoH domains (HCAU, ES, SCC, NBE) using Confirmatory Factor Analysis (CFA) (Figure 1). Education only had one measure so was maintained as an individual-level variable. This approach allows for estimation of latent constructs consistent with the HP2030 framework and addresses multicollinearity concerns stemming from modest intercorrelations among survey items within domains (Supplementary Figure 3). Most SDoH domain models demonstrated good model fit, with the exception of the overall SDoH model, which had an acceptable CFI of 0.92 (Supplementary Table 2).

The latent SDoH construct was most strongly explained by the Economic Stability (ES) domain (factor loading (λ) = 0.93), followed by the HCAU (λ = 0.89) and Social and Community Context (SCC; λ = 0.86) domains. Within ES, food insecurity emerged as the most prominent indicator (λ = -0.66). For HCAU, key drivers included difficulty affording care (λ = -0.57), concerns about medical costs (λ = -0.59), and delays in receiving care (λ = -0.61; e.g., due to financial constraints, being unable to get off of work, or discomfort with the healthcare system). In the SCC domain, perceived stress and loneliness showed the strongest factor loadings(λ = -0.76, λ = -0.74, respectively), followed by everyday discrimination (λ = -0.62). The distributions of these normalized SDoH domains in the direction of increased social risk are available in Supplementary Figure 4.

### Evaluation of the Association between SDoH Domains vs. Component Measures with Disease Prevalence

To better understand how SDoH affects human health, we evaluated the strength of the association between latent SDoH constructs and nine chronic conditions, comparing them to the predictive performance of their individual survey components. Each model was adjusted for age, age^2^, Sex/Gender, record depth, visit frequency and tested for the strength of association between a single SDoH variable and disease outcome within the Individual Cohort. Effect sizes represent the change in disease liability (z-score) per one-unit increase in the predictor—corresponding to the difference between the lowest and highest observed values within each domain.

Overall, latent SDoH constructs demonstrated stronger and more consistent associations with disease outcomes compared to individual items, as reflected in higher median effect sizes across conditions (Figure 2). Overlapping confidence intervals suggest that specific components may, in some cases and for some diseases, perform as well or better than their broader domain construct. For example, the individual measure of income as a percentage of the poverty threshold (“percent of poverty threshold”) outperformed the ES construct in predicting Type 1 Diabetes (T1D), with a larger absolute effect size of 1.16 [0.97, 1.34] compared to 1.01 [0.81, 1.20] for ES, although the error bars overlap (Supplementary Figure 5; Supplementary Table 3 S1). Moreover, although SCC is not significantly associated with Breast Cancer, “Social Cohesion” and “Social Support” are.

**Figure 2:**
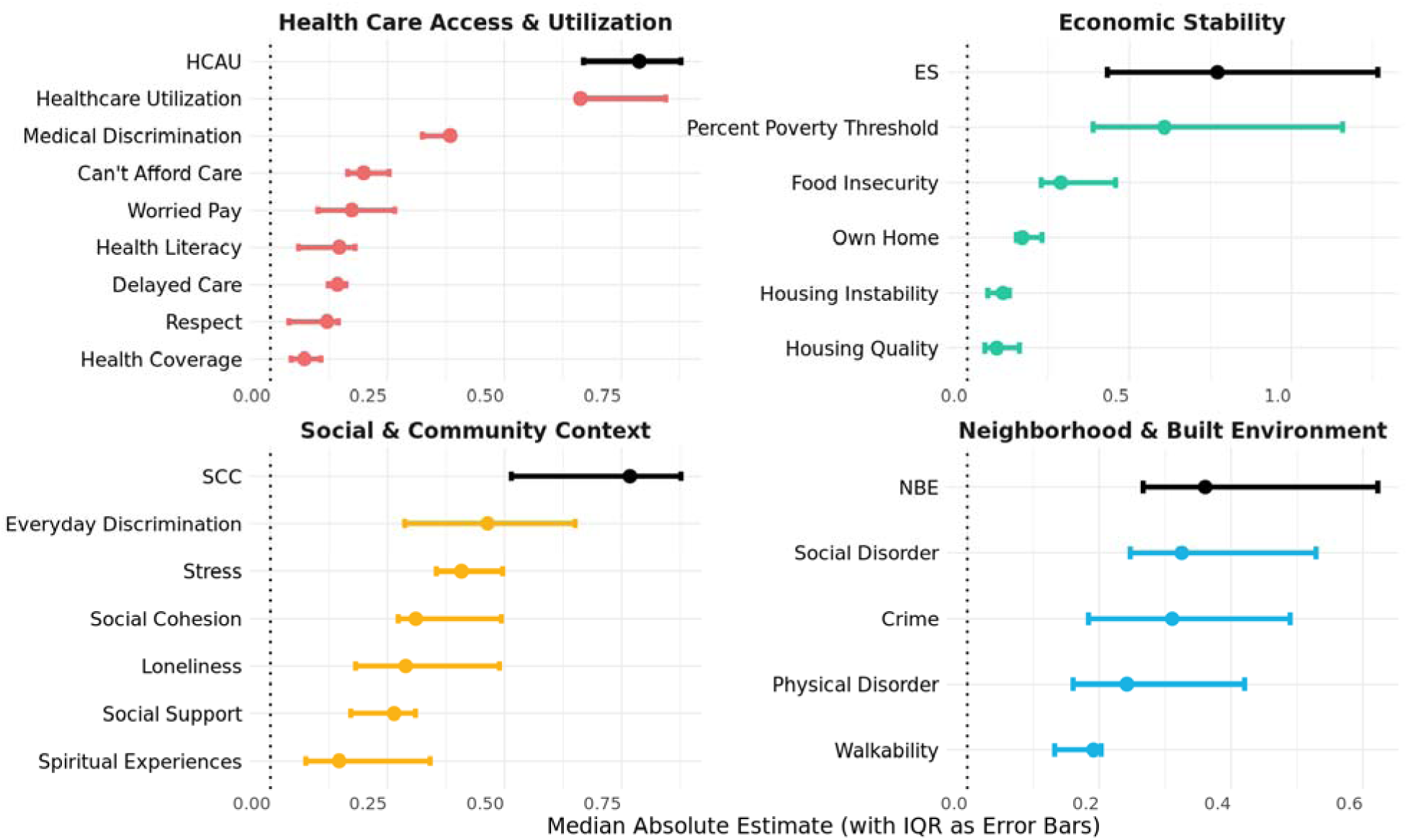
Latent SDoH Domain Constructs Outperform Components Across Nine Chronic Conditions. Estimates from SEMs assess the association between each SDoH domain and their components with nine chronic conditions. The y-axis is ordered by the median estimate across the conditions, with interquartile ranges (IQRs) represented as error bars. Component items are color-coded by domain, while latent constructs are depicted in black.

### Evaluation of the Association between Individual- vs. Area-level SDoH Measures with Disease Prevalence

To evaluate the relative predictive value of individual v. area-level measures of SDoH, we compared median effect sizes across nine chronic conditions in the Individual Cohort. Overall, we observed no consistent advantage of individual-level versus area-level SDoH measures in predicting disease status (Figure 3A; Supplementary Table 3 S2). Instead, the two sets of metrics appear to capture distinct dimensions of social determinants, as evidenced by their limited correlation (Figure 3B).

**Figure 3:**
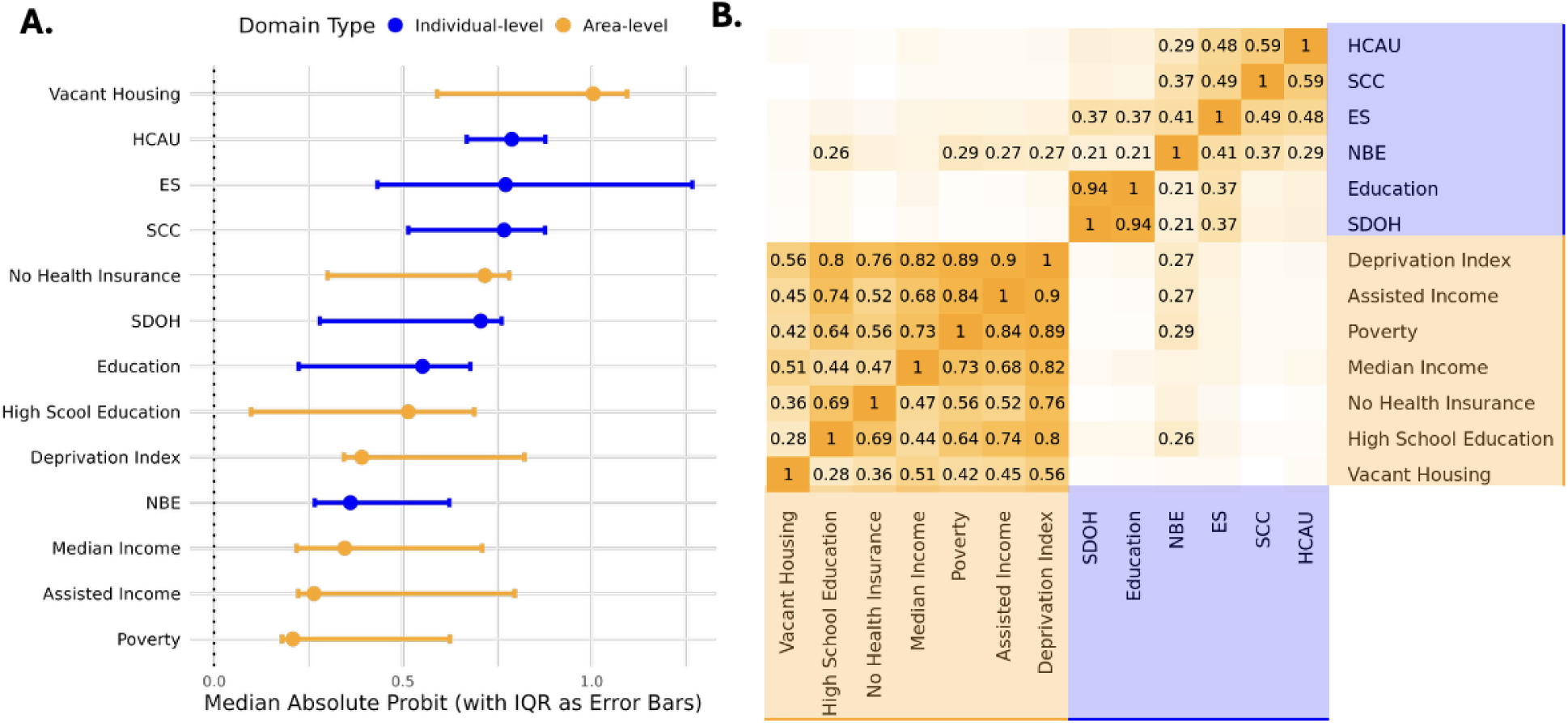
Comparison of Individual-Level and Area-Level SDoH. **A.** Probit coefficients from SEM models assessing the association between each SDoH domain and disease status. Models are adjusted for age, age^2^, Sex/Gender, record depth, and visit frequency. The y-axis is ordered by the median absolute estimate across nine chronic conditions, with interquartile ranges (IQRs) represented as error bars. Individual-level metrics are depicted in blue; area-level metrics are shown in yellow. **B.** Correlation heatmap illustrating relationships between individual-level and area-level SDoH metrics that were min-max normalized to be in the direction of risk (i.e. percent with high school education and median income were flipped). Area-level metrics are highlighted in orange and individual-level metrics are highlighted in blue. Only correlations exceeding 0.2 are annotated with their respective values.

The area-level metric percentage of vacant housing emerged as the strongest predictor of disease status overall (median absolute probit effect size 1.00 [IQR: 0.59, 1.09]), followed by the individual-level measure of HCAU (0.79 [.67, .88]). NBE is the only individual-level metric modestly correlated (range: r=0.8-0.29) with area-level metrics, indicating that it may serve as a conceptual and empirical bridge between the two levels of measurement. Interestingly, Education exhibits a strong correlation with the overall SDoH metric (r=0.94), despite having the lowest factor loading (λ = 0.28).

To evaluate the influence of SDoH on nine chronic conditions, we analyzed estimates from SEM models for both individual-level and area-level data in the Individual Cohort. SEM was used to control for the correlation between SDoH metrics and covariates included in the analysis (age, age^2^, Sex/Gender, record depth, and visit frequency) that also have significant impacts on disease status (Supplementary Figure 3). Distinct patterns of association emerged at each level, suggesting that individual- and area-level measures capture different aspects of the social environment (Figure 4). For example, while prostate cancer is inversely associated with individual-level SDoH metrics, this is not true for all area-level measures.

**Figure 4:**
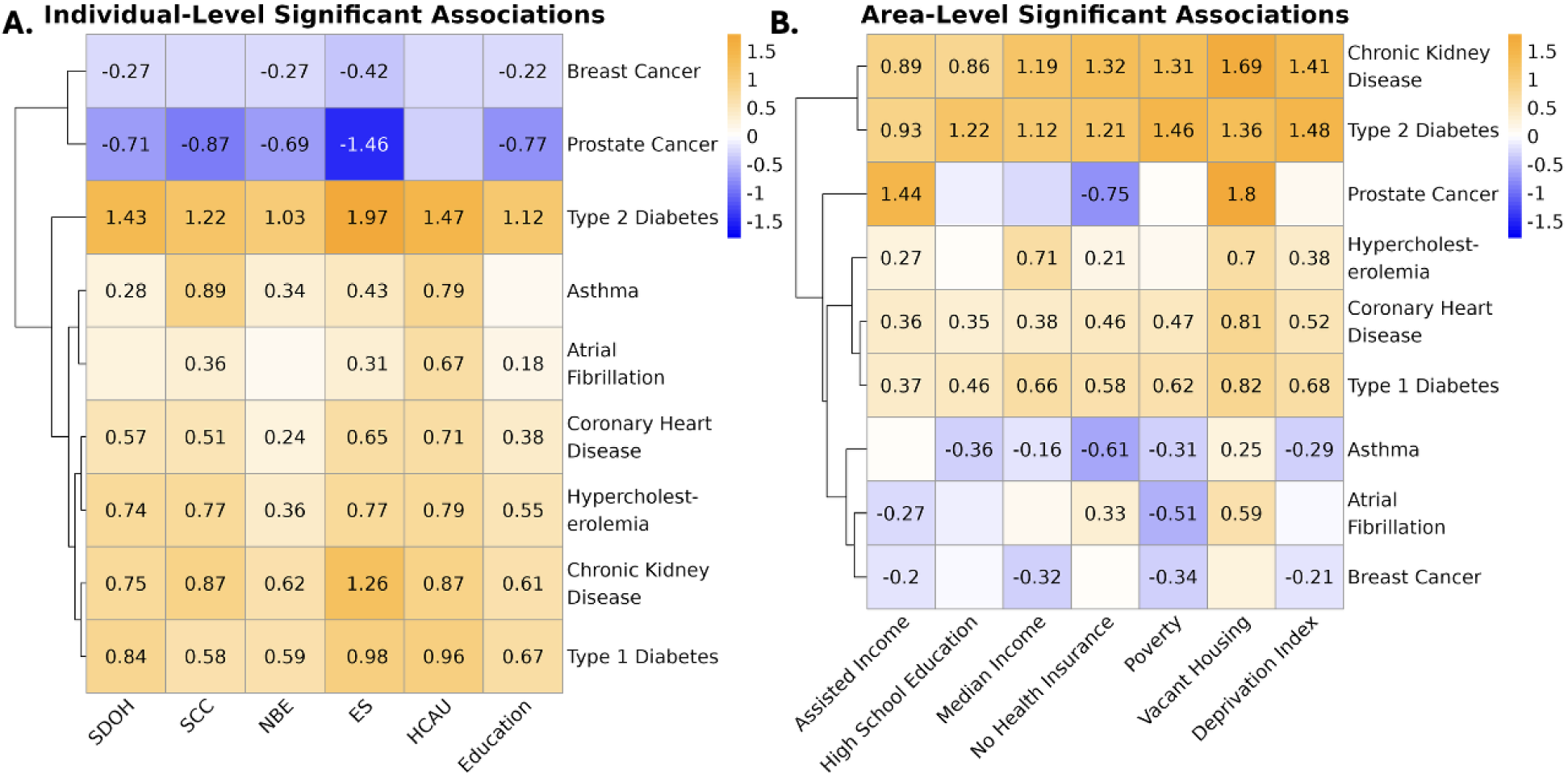
Distinct Patterns of Associations Between SDoH and Nine Chronic Conditions at Individual and Area Levels. **A.** Heatmap displaying the estimates from SEMs assessing the relationship between individual-level SDoH domains and nine chronic conditions in the Individual Cohort. Only statistically significant associations are annotated with effect sizes. Negative values indicate that higher levels in a specific SDoH risk are associated with reduced probability of disease. **B.** Heatmap illustrating the probit estimates from SEMs evaluating area-level SDoH domains in the SES Cohort. For both heatmaps, negative values indicate that higher levels in a specific SDoH risk are associated with reduced probability of disease. Both heatmaps are on the same scale to facilitate direct comparison between individual-level and area-level SDoH associations, though the order of disease outcomes varies.

T1D, T2D, chronic kidney disease (CKD), and asthma appear to be most strongly associated with adverse individual-level SDoH. Notably, breast cancer and prostate cancer exhibit opposite directions of association from the rest of the chronic conditions, and Afib has few significant associations. The composite SDOH score does not outperform individual domains for most conditions. In fact, each condition exhibits a unique pattern of association with individual-level SDOH metrics. While no domain consistently outperforms the others for all diseases, ES (abs average probit effect size: .92) and HCAU (.78) tended to be the strongest predictors overall (Figure 4A).

Different trends are seen at the area-level, where T1D, T2D, coronary heart disease (CHD), and CKD appear to be the most strongly influenced by area-level SDoH. At the area-level, breast cancer and prostate cancer exhibit opposite directions of effects for some indices but not all. Similarly to individual-level findings, the composite deprivation index (NCDI) does not have a stronger magnitude of effect than the more specific indices. Area-level rates of vacant housing is the strongest predictor overall (abs average: .91), and maintains its direction of effect across all diseases (Figure 4B). While asthma and Afib maintain the same direction of effect as expected at the individual-level, with increasing social risk leading to increased disease prevalence, this is not the case for all area-level metrics. For instance, at the 3-digit zip code level, adults living in regions with lower proportions of adults with a high school education, health insurance coverage, and poverty have a higher risk of asthma.

Moreover, in stratified analyses, we found that the associations between SDoH and disease can vary across SIRE groups (Supplementary Figure 6; Supplementary Table 3 S3). For example,the magnitude of the association between vacant housing and prostate cancer in NHB (2.98 [95% CI: 1.97, 4.00]) and HS (3.08 [2.00, 4.15]) populations is double the association found in the NHW group (1.45 [1.16, 1.74]). We even observe a change in the direction of effect of percent receiving assisted income on T2D between NHW (0.45 [0.33-0.58]) and HS (-0.61 [-0.79, -0.44] populations. Using Cochran’s Q test, 40 of 81 disease-trait pairs have significant heterogeneity across SIRE groups at the Bonferroni corrected significance threshold of 6.17E-04 (.05/81), which may be in part due to underlying differences in SDoH distributions across these populations, but could also be due to group-specific dynamics with structural factors and their respective access to those factors (Supplementary Figure 7).

### Disease-specific polysocial risk prediction models incorporating individual- and area-level metrics match or outperform SIRE in disease prediction modeling

Given the disease-specific patterns of associations with SDoH, we used elastic net regression to select variables from correlated items and generate disease-specific polysocial risk scores (PsRS) (Figure 5; Supplementary Table 4 S1:3). For some traits such as CHD, breast cancer, and hypercholesterolemia, including SDoH metrics beyond record depth and visit frequency (EHR model) did not show much improvement in model performance compared to the Base model. We also observed that, in most cases, including all five HP2030 domains in the model (Domain PsRS model) did not significantly outperform models using just income and education (SES model) in the Individual-SIRE Cohort; however, including domain components in addition to summary domains (Component PsRS model) did offer statistically significant improvements for some traits such as T1D (AUC of 0.73 [95% CI: 0.72,0.75] for the Component PsRS model vs 0.70 [0.68,0.73] for the SES model (p=4.44E-06)). Alternatively, including income and education as separate variables in the model (SES model) led to better performance than the Component PsRS model for some traits such as CHD (p=8.54E-07) and CKD (p=6.27E-12).

**Figure 5:**
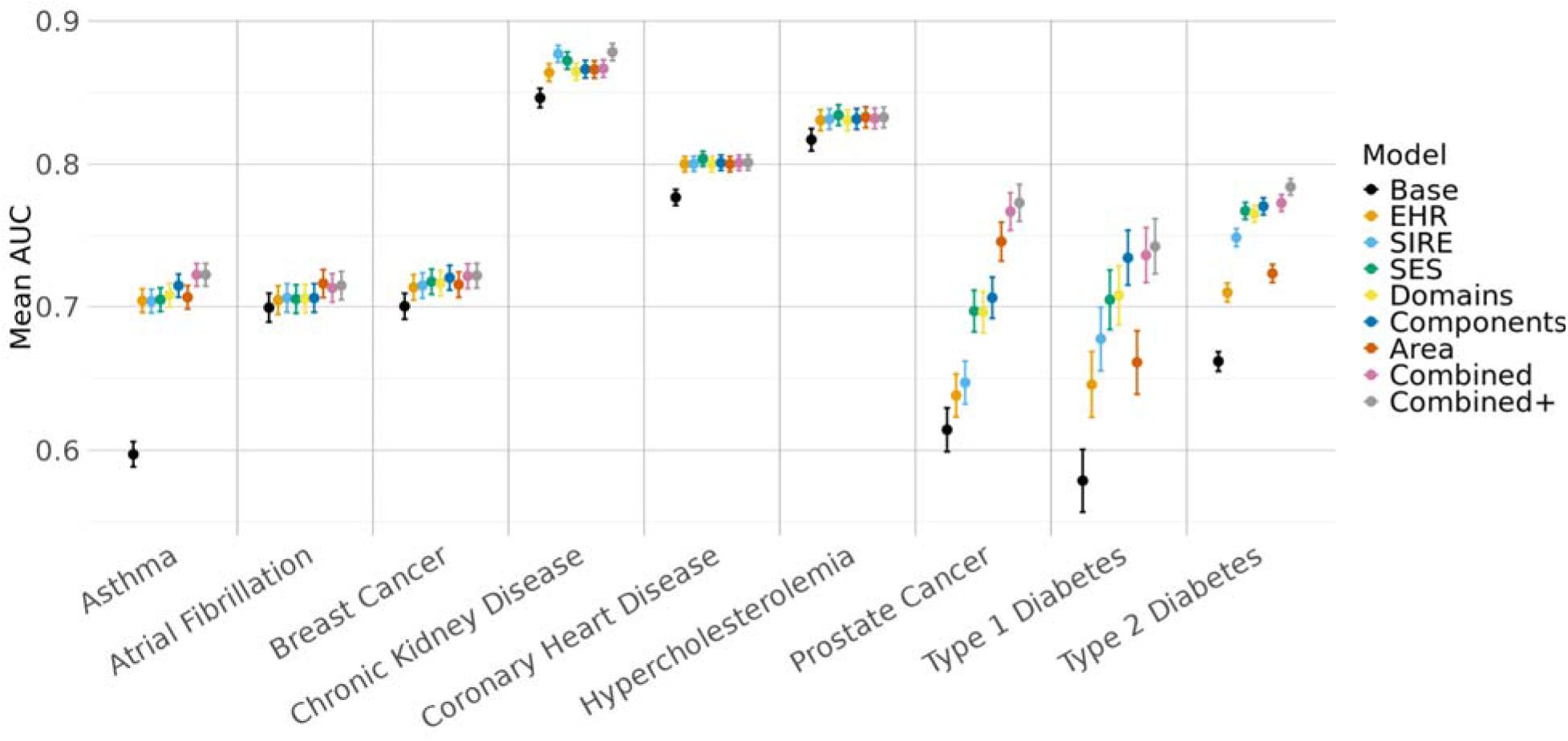
Evaluation of Prediction Models Incorporating Various Levels of SDoH and SIRE in the Individual Cohort. Dot plot showing the mean AUC and corresponding 95% confidence intervals from 5-fold cross-validated elastic net models using a 70/30 training/testing split. All models were run in the Individual Cohort. The x-axis is organized by disease. Each dot is color coded by model type. The Base model includes age, age^2^, and Sex/Gender. In addition to the covariates included in the Base model, the EHR model also includes visit frequency and record depth. The rest of the models build on the EHR mode. The SIRE model also includes NHB and HS. The SES model also includes percent of poverty threshold and education. The Domain model also includes a PsRS constructed from the SDoH domains and the Component model additionally includes the components used in the construction of the domains. The Area model includes the PsRS trained on all area-level metrics. The Combined model includes a PsRS trained on the domains, components, and area-level metrics and the Combined+ model additionally includes NHB and HS.

Diseases also showed variation in whether individual-level or area-level metrics are more predictive. For example, prostate cancer models had better prediction from area-level metrics (Area PsRS model) with an AUC of 0.75 [95% CI: 0.73,0.76] compared to individual-level metrics (Component PsRS model; 0.71 [0.69, 0.72]; p=3.94E-07). On the other hand, Component PsRS models outperform Area models for diabetes - 0.73 [0.72,0.75] vs 0.66 [0.64,0.68] for T1D (p=2.05E-12) and 0.77 [0.76,0.78] vs 0.72 [0.72,0.73] (p=8.58E-73) for T2D, respectively. Moreover, combining individual-level and area-level metrics into a single model (Combined PsRS model), led to significant improvements for asthma, T2D, and prostate cancer, relative to either model alone.

SIRE can correlate strongly with SDoH, which is a large part of why it has historically been used in disease modeling (Supplementary Figure 8). However, we find that the Combined SDoH-based PsRS model, which does not include race or ethnicity, matches or outperforms the SIRE model across nearly all traits. For instance, the Combined PsRS model (0.77 [0.77,0.78]) outperforms the SIRE model (0.74 [0.74,0.75]) in predicting T2D (p=1.37E-20). The only exception is CKD (0.87 [0.86,0.87] for the Combined PsRS model vs 0.88 [0.87,0.88] for the SIRE model (p=2.34E-15)), which may be partly attributable to the historical incorporation of race into estimated glomerular filtration rate (eGFR) calculations and CKD diagnostic practices ^29^. Additionally, adding SIRE to the Combined PsRS model (Combined PsRS+ model) yielded additional improvements for CKD and diabetes. These trends were largely consistent in the more socioeconomically and demographically diverse SES-SIRE Cohort, with some loss of predictive accuracy in the Combined PsRS model for T1D and asthma without the richer individual-level data (Supplementary Figure 9; Supplementary Table 4 S4:6).

Due to the previously observed difference in patterns of association of some SDoH across SIRE, we also ran analyses stratified by SIRE (Supplementary Figure 10; Supplementary Table 4 S7:9). This analysis reveals some distinct patterns across socially defined groups. For example, SDoH improve prediction of asthma only in the NHW population. Additionally, the SDoH model adds predictive capability for T2D in HS (0.83 [0.82,0.84] vs 0.79 [0.78,0.80] (p=1.70E-33)) and NHW (0.76 [0.76,0.77] vs 0.72 [0.72,0.73] (p=1.15E-110)) populations, but not the NHB population (0.75 [0.74,0.76] vs 0.75 [0.74,0.76] (p=0.47)) compared to the EHR model. The same pattern is observed for CKD and can likely be explained by the narrow distribution of the Combined PsRS in the NHB population for CKD and T2D (Supplementary Figure 11).

### Targeted Phenome-Wide Association Study

Interested in determining if incorporating SDoH into disease model prediction may be of interest to a broader range of traits, we conducted a targeted Phenome-Wide Association Study looking at the broader disease groupings of the nine chronic conditions explored in depth above (Figure 6; Supplementary Table 5 S1). Across all five disease groups and 513 phecodes, approximately 58% of phecodes showed significant associations with at least one SDoH in the Individual Cohort after adjusting for age at most recent phecode, Sex/Gender, record depth, and visit frequency. This proportion was lowest for neoplasms (37%) and highest for respiratory disorders (67%) and endocrine/metabolic disorders (67%) (Supplementary Table 5). Notably, when stratifying associations by directionality, neoplasms exhibited a distinct pattern, with many effects in the opposite direction compared to other groups, especially for individual-level SDoH metrics (Figure 6A). Additionally, several phecodes - including renal failure, hypertension, and congestive heart failure-were significantly associated with 10 or more SDoH (Figure 6B).

**Figure 6:**
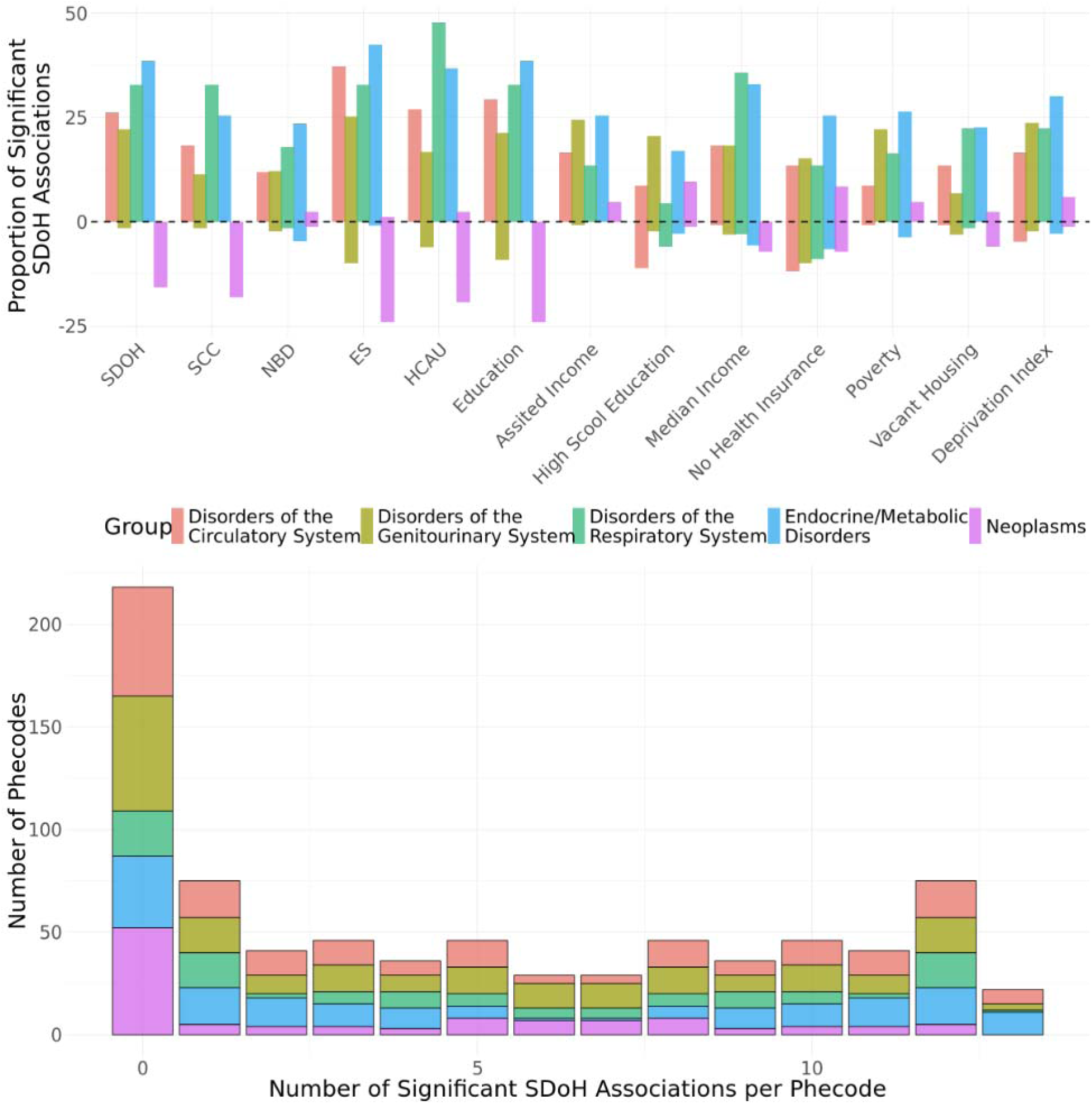
Targeted Phenome-Wide Association Study of SDOH in Individual Cohort. **A**. Box plot showing the proportion of significant SDoH associations for each disease group. The y-axis represents the proportion of significant associations, split into positive (above the x-axis, associated with increased disease prevalence) and negative (below the x-axis, associated with decreased disease prevalence) effects. Boxes are grouped by SDoH category and color-coded by disease group. **B.** Histogram showing the distribution of significant SDoH associations across phecodes. The x-axis indicates the number of SDoH significantly associated with each phecode (maximum SDoH tested = 13), while the y-axis shows the count of phecodes with that number of associations. The colors correspond to the same group colors in panel A.

We repeated this analysis in the SES Cohort, which has an increased sample size, allowing for the investigation of associations with more traits that meet the case threshold of 100 (Supplementary Figure 12; Supplementary Table 5 S2). In the SES Cohort, 69% of phecodes (N=621) showed significant associations with at least one SDoH (Table 2). This proportion remained lowest for neoplasms (44%) and was highest for respiratory disorders (83%). The inverse associations with neoplasms remained in the SES Cohort (Supplementary Figure 12). Associations with “No Health Insurance” also appeared to have less consistent directions of effects than other measures in all cohorts (Figure 6A; Supplementary Figure 12A).

**Table 2:** Proportion of Significant Phecodes in Targeted PheWAS by Cohort.

| **Group** | **Individual Cohort**<br>**(n=54,313)** | **SES Cohort**<br>**(n=125,295)** |
| --- | --- | --- |
| Circulatory system | 73 / 126 (57.94) | 107 / 151 (70.86) |
| Endocrine/ metabolic | 71 / 106 (66.98) | 96 / 129 (74.42) |
| Genitourinary | 75 / 131 (57.25) | 112 / 154 (72.73) |
| Neoplasms | 31 / 83 (37.35) | 48 / 108 (44.44) |
| Respiratory | 45 / 67 (67.16) | 65 / 79 (82.82) |
| Total: | 295 / 513 (57.50) | 428 / 621 (68.92) |

## Discussion

This study demonstrates the power and complexity of incorporating individual-level and area-level SDoH into disease prediction models using the expansive data from the All of Us Research Program. By applying CFA, we aggregated related individual SDoH measures into latent domains aligned with the Healthy People 2030 framework, enabling dimensionality reduction while preserving conceptual clarity and enhancing generalizability across cohorts with related measures. Our results reveal that diseases have distinct “social architectures,” where the predictive strength, the most relevant SDoH domains, and the relative contributions of individual-versus area-level SDoH vary by condition. This prompted the development of disease-specific PsRS. Across models, we found that the individual-level variables of record depth, visit frequency, income, and education captured most of the predictive power of individual-level SDoH for some traits. Many conditions showed improved performance when both individual- and area-level data were included, where combined models matched or outperformed models using SIRE. Moreover, we find that SDoH may have varying impact in predicting disease case control status across populations delineated by SIRE, further complicating the relationship between SDoH and disease in diverse populations. Our PheWAS suggests value in future work around investigating the utility of including SDoH in clinical risk estimation for common complex diseases. This work highlights the value of including SDoH in models to enhance disease prediction and serve as more precise, interpretable, and intervenable alternatives to SIRE in disease modeling, while pointing to the need of future work investigating the intersectionality of these measures in disease.

While we initially set out to identify universal SDoH predictors of disease—including constructing a composite individual-level SDoH metric—we found that each disease, as captured by the healthcare systems included in All of Us, exhibits a distinct “social architecture,” differing in which measures, at which levels of measurement, and to what degree the SDoH associated with each disease. For instance, CKD and T2D were most strongly associated with economic stability and percent of poverty threshold, whereas asthma and Afib were more strongly linked to HCAU. To account for these disease-specific patterns, we developed tailored PsRS for each condition using elastic net models to select relative parameters and shrink estimates from related measures. For some diseases, such as breast cancer, CHD, and hypercholesterolemia, while incorporating visit frequency and record depth improved model performance, adding SDoH data beyond these factors offered little additional predictive value, suggesting either that HCAU alone contributes to recorded disease prevalence or that other predictive SDoH (e.g., ES) are closely interrelated with visit frequency and record depth. Prostate cancer had a substantial increase in model performance when including area-level SDoH metrics, while individual-level data were more informative for conditions such as T1D and T2D. These findings may reflect disease etiology in which T2D is more strongly influenced by lifestyle factors, where access to healthy foods is very important and driven by ES, whereas prostate cancer is less strongly affected by lifestyle and is often detected incidentally through routine screening, making area-level healthcare access a potentially greater determinant of diagnosis.

Since individual-level SDoH data did not consistently outperform area-level measures in predicting disease outcomes—and given that these data types appear to capture distinct aspects of the social environment—we evaluated whether combining both sources in a single model would enhance disease prediction. This approach significantly improved disease prediction models for asthma, T2D, prostate cancer. Moreover, combined SDoH models matched or outperformed SIRE models, suggesting that SDoH may better capture the underlying processes and serve as a more interpretable option. However, the effects of the combined SDoH models were not consistent across SIRE populations. For example, the SDoH metrics available in AoU are not predictive of asthma in NHB or HS populations, or of T2D or CKD in NHB populations, despite being predictive in other SIRE groups. Additionally, some SDoH measures have up to a two-fold difference in effect size or reversed directions of effect between SIRE groups.

Differences in effects may in part be due to differences in distributions of SDoH across these populations.Given this population-specific intersectionality, survey response bias may bias our disease-specific PsRS such that predictive accuracy is greater in socially advantaged and NHW groups, the largest group with the most diverse SDoH, emphasizing the importance of diverse and representative cohorts for training these scores. For example, discrimination may hold a heavier weight in PsRS if the model was trained in a more racially diverse group. Moreover, some models showed a gain in predictive performance when SDoH metrics were combined with SIRE, which may further point to intersectionality of these measures across populations or to the SDoH survey not fully capturing determinants of structural inequality. As another example, Marginalization-related Diminished Returns (MDRs) - the phenomenon in which high SES returns smaller health benefits in marginalized populations - may affect model accuracy across SIRE groups. For example, education reduces the risk of heart disease more strongly for a NHW population than for HS or Black populations ^30^. Therefore, future research should investigate how SDoH may influence disease risk differentially across populations and prioritize inclusion of diverse participants across SIRE groups.

Use of area-level indicators may offer unique advantages in terms of broader sample diversity and representativeness, facilitating reproducibility across studies and populations, and capturing factors that affect health across a population; however, they may not be enough to fully capture the impact of SDoH on disease. Moreover, these metrics are likely to provide limited additional value in studies with narrow geographic coverage. Ultimately, future studies will need to consider the trade-offs of the richness of individual-level data with the increased representativeness and harmonizability of area-level metrics, depending on research goals, the trait of interest, and data availability. At the time of V8 in the AoU cohort, using more broadly available income and education metrics alongside area-level metrics may offer the best trade-off of these advantages for predicting case-control status until the SDoH survey is more widely administered and captures a more diverse group of study participants. The value of these metrics, however, depends on study goals and outcomes; for example, T1D and asthma prediction improve significantly with SDoH survey data. Use of individual-level data may also improve interpretability or reveal targets for intervention. Moreover, similar metrics are needed across studies in order to facilitate cross-study validation and test the portability of disease-specific PsRS.

Using an EHR-based cohort to investigate the effects of SDoH on disease introduces real-world biases that may affect disease capture. Importantly, rather than capturing real disease prevalence, our results reflect associations with observable diagnosis in EHR records which are affected by SDoH such as healthcare access, visit frequency, and provider biases, as well as differences in EHR completeness in the data set. This can be observed in the heterogeneous effects of “No Health Insurance” on disease prevalence in the PheWAS: while not having health insurance may increase disease prevalence it will also reduce disease capture. This phenomena will especially affect diseases with prolonged asymptomatic periods and may introduce collider bias on all SDoH ^31^. For example, we found counterintuitive directions of association between individual-level SDoH and breast cancer and prostate cancer, wherein more socially advantaged groups had higher probability of disease. This effect was mirrored and extended in the PheWAS, in which the direction of effects for diseases in the neoplasm group was often flipped. Increased screening in higher SES populations may detect indolent and subclinical forms of cancer and result in overdiagnosis ^32^. On the other hand, racial disparities in cancer outcomes are driven in part by later diagnosis, where racialized groups are more likely to have advanced stage or metastatic cancer at diagnosis ^33–37^. Further, redlining and neighborhood measures of segregation have been associated with later stage cancer at presentation, possibly explained by reduced cancer screening ^38,39^. Taken together, these findings point to the need to increase cancer screenings in low SES and historically redlined areas to address disparities in early detection and outcomes ^40,41^. The inverse associations of SDoH with neoplasms also indicates that these real-world biases may be decreasing the strength of associations between SDoH and disease, as well as SIRE and disease. These biases may also explain the lack of association between many of the SDoH variables and Afib, which may also have a prolonged asymptomatic phase requiring specific screening for diagnosis. These real-world biases and limitations underscore the importance of using traditional population-based research cohorts (as well as cohorts in areas of the world with universal healthcare) with careful study design in their health monitoring to better understand the impact of SDoH on disease (given more uniform screening for diagnoses such as Afib).

While AoU offers the richest individual-level SDoH data, and in the largest population, of any national health-related study within the US to our knowledge, there are still some limitations with this data. Most notably, there is significant non-random missingness (>90% with insufficient SDoH survey completion) which reduces the size and representativeness of the cohort, introduces selection bias, and may limit the generalizability of findings. Additionally, recruitment bias may be leading to different distributions of SDoH within socially defined groups, such as the relatively narrower distribution of SDoH in the NHB population. Further, while SDoH such as educational attainment are generally stable throughout the adult lifespan after a certain age, AoU currently only offers a single timepoint for both individual- and area-level metrics.

Moreover, the surveys assay adult measures of SDoH; however, childhood and early life measures, while correlated with adult measures, may capture different and earlier SDoH exposures important to the causal framework and may be more important in determining disease risk later in life. For instance, the social vulnerability index has been found to associate with risk of obesity most strongly at birth compared to later life stages ^42^. Additionally, the All of Us area-level data is currently encoded at the 3-digit zip code level to protect participant privacy, limiting granularity of area-level measures. Large heterogeneity could be present at this spatial scale, and 3-digit zip could be serving as a proxy for other factors such as urbanicity, geography (i.e. higher social deprivation on average in the South), or even segregation. Moreover, use of area-level data to inform patient-level interventions is subject to misinterpretation such as the ecological fallacy in which incorrect assumptions are made at the individual-level based on a group-level measure ^43^.

It is important to note that we cannot claim causality. Our analyses use disease prevalence rather than incidence due to the low number of incident cases following survey completion in AoU. As a result, we cannot establish temporal ordering in which the exposure proceeds the outcome. For instance, while we observed an association between higher levels of spirituality and diabetes, having diabetes could be leading to increased spirituality, or a third variable, such as cultural practices, could be confounding our SDoH exposure and our disease outcome ^44^. As incident cases increase in All of Us, future studies should focus on the association of SDoH with incident disease to establish temporal ordering. Moreover, we analyzed associations with prevalent disease, but SDoH may be more strongly associated with stage at diagnosis, treatment quality, hospitalization, and disease outcomes, depending on the disease studied ^45–50^. Future research should also investigate the specific pathways through which SDoH mediate disease risk, such as limited access to healthy foods or safe environments for physical activity, health literacy, childhood adversity, chronic stress, and exposure to environmental toxins ^51–53^. Further work is also needed on how SDoH can inform care, such as increased training for nurses, integrating social or community health workers into healthcare systems, or direct intervention on health-related social needs by providers or payers ^6,54–56^.

Our results demonstrate that diseases have unique “social architectures,” prompting the development of disease-specific PsRS. Because individual-level and area-level SDoH are weakly correlated, they provide distinct signals that can enhance PsRS. Given the strong selection bias in the individual-level SDoH surveys, use of individual-level education and income variables may offer the best trade-off between granularity and increased representativeness and harmonizability for some traits; however, some traits showed significant improvement when the full set of individual-level SDoH survey measures was included. Ultimately, the choice of SDoH variables should be made on a trait-by-trait basis. While PsRS matched or outperformed SIRE-based models, the effects of SDoH varied across SIRE groups, highlighting the need for population-specific considerations and a diverse training sample. Our PheWAS demonstrated that many outcomes could likely benefit from the incorporation of SDoH into disease prediction modeling, allowing a shift in focus from group labels and individual behaviors to structural drivers of health ^25^. Such insights can inform policy changes—such as Medicaid expansion—that improve healthcare access and reduce chronic disease burden ^57^. Ultimately, strengthening shared resources, social capital, and partnerships among hospitals, public health agencies, and social services will be critical for improving population health, narrowing health disparities, and extending life expectancy.

## Supporting information

Title Page

## Data Availability

All data produced are available online as a community workspace on the All of Us platform. The name of the workspace is "Social Determinants of Health Measurement Models V8"

## Acknowledgments

The authors thank the participants and research teams from the All of Us research program, without whom this research would not have been possible. This research was conducted with support and resources provided by the Odum Institute for Research in Social Science at UNC-Chapel Hill. We would specifically like to thank Chris Wiesen for providing his expertise. We would also like to thank the Polygenic Risk Methods Development (PRIMED) Consortium Social Determinants of Health Working Group. A list of individuals included in PRIMED consortium banner are at https://primedconsortium.org/publications/banner. We also thank the Diabetes Algorithm contributors: Katie Taylor, Josep Mercader, Alisa Manning, Alicia Huerta-Chagoya, Raymond Kreienkamp, Maheak Vora, Ravi Mandla, Sara Cromer, Kaavya Ashok, Aaron Deutsch.

## Funding sources

Research reported in this publication was supported by the National Institutes of Health for the project “Polygenic Risk Methods Development (PRIMED) Consortium”, with grant funding for study sites D-PRISM (U01HG011723), EPIC-PRS (U01HG011720), CAPE (U01HG011715), and the coordinating center (U01HG011697). The content is solely the responsibility of the authors and does not necessarily represent the official views of the National Institutes of Health. SJC was supported by the American Diabetes Association (7-21-JDFM-005). GLW was supported by the NHGRI (R35HG011944). MG was supported by the National Institute On Aging of the National Institutes of Health (F99AG088695).

## Notes

### Competing Interest Statement

SJC is Ad hoc consulting for Alexion Pharmaceuticals and Patient Square Capital and their spouse works for Depuy-Synthes. IRK received an honorarium from Sage Bionetworks during the past 12 months

### Author Declarations

the All of Us Research Program

