## Supplementary pdf for "Quantifying Social Determinants of Health for Disease Prediction: A Multi-Level Approach Using Healthy People 2030 and All of Us Data"

### Supplementary Tables

#### Supplementary Table 1

**Supplementary Table 1 (S1):** Area-level SDoH across all cohorts

**Supplementary Table 1 (S2):** Demographic data by SIRE across all cohorts

**Supplementary Table 1 (S3):** Case control demographics in Individual Cohort

**Supplementary Table 1 (S4):** Case control demographics in SES Cohort

#### Supplementary Table 2

**Supplementary Table 2 (S1):** CFA model fit statistics for each SDoH domain and composite SDoH measure.

**Supplementary Table 2 (S2):** CFA model parameter estimates for ES

**Supplementary Table 2 (S3):** CFA model parameter estimates for HCAU

**Supplementary Table 2 (S4):** CFA model parameter estimates for NBD

**Supplementary Table 2 (S5):** CFA model parameter estimates for SCC

**Supplementary Table 2 (S6):** CFA model parameter estimates for SDOH

#### Supplementary Table 3

**Supplementary Table 3 (S1):** Disease SDOH association effect sizes from SEM

**Supplementary Table 3 (S2):** Median of SDOH association effect sizes across diseases

**Supplementary Table 3 (S3):** Disease SDOH association effect sizes in stratified analysis with heterogeneity statistics

#### Supplementary Table 4

**Supplementary Table 4 (S1):** PsRS EN summary results in Individual cohort

**Supplementary Table 4 (S2):** Logistic regression summary results in Individual cohort

**Supplementary Table 4 (S3):** AUC pair-wise comparisons in Individual cohort

**Supplementary Table 4 (S4):** PsRS EN summary results in SES cohort

**Supplementary Table 4 (S5):** Logistic regression summary results in SES cohort

**Supplementary Table 4 (S6):** AUC pair-wise comparisons in SES cohort

**Supplementary Table 4 (S7):** Stratified EN summary results in SES cohort

**Supplementary Table 4 (S8):** Stratified logistic regression summary results in SES cohort

**Supplementary Table 4 (S9):** AUC pair-wise comparisons of stratified results

#### Supplementary Table 5

**Supplementary Table 5 (S1):** PheWAS results in Individual Cohort across 13 predictors

**Supplementary Table 5 (S2):** PheWAS results in SES Cohort across 9 predictors

### Supplementary Figures

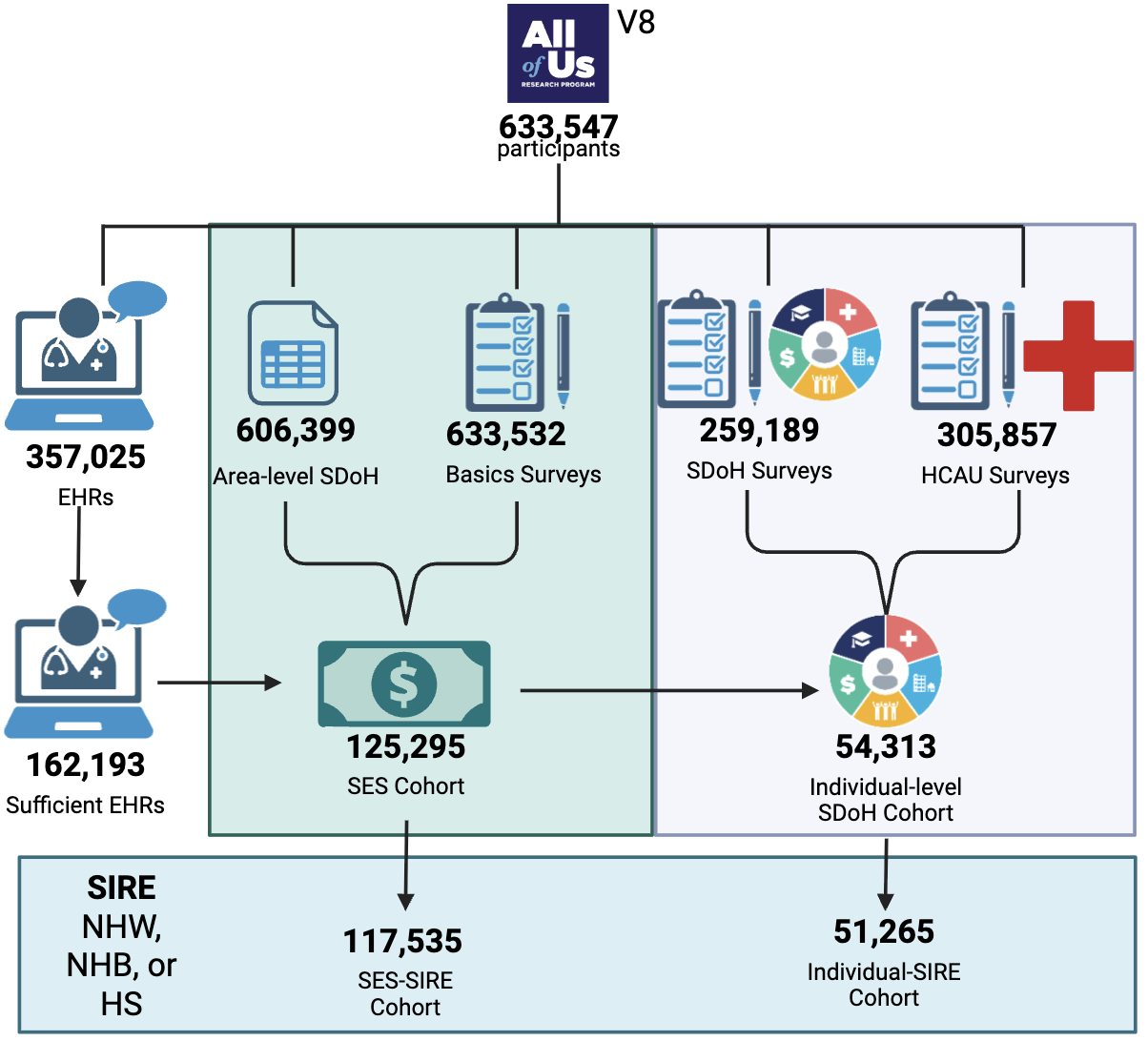

**Figure 1: Study Participant Flow Chart**

Version Release 8 (V8) of the All of Us Research Program contains data on 633,547 participants. Of those, 357,025 have linked Electronic Health Records (EHRs) and 162,193 meet the criteria for being “sufficient” based on having at least three entries and being at least three years long to meet “medical home” definition. 633,537, 259,189, and 305,857 participants filled out the basics, SDoH, and HCAU surveys respectively. Income data was divided by the corresponding poverty threshold based on location, size of household, and month/year of survey to get the percentage of the poverty threshold. 125,295 individuals had this data in addition to education to make up the SES cohort. Of these, 54,697 filled out the Individual-Level SDoH survey with at least 60% completeness for each domain. Of the Individual and SES Cohorts, 51,265 and 117,535 self-identified as non-Hispanic White (NHW), non-Hispanic Black (NHB), or Hispanic (HS),respectively, to make up the Individual-SIRE and SES-SIRE subcohorts.

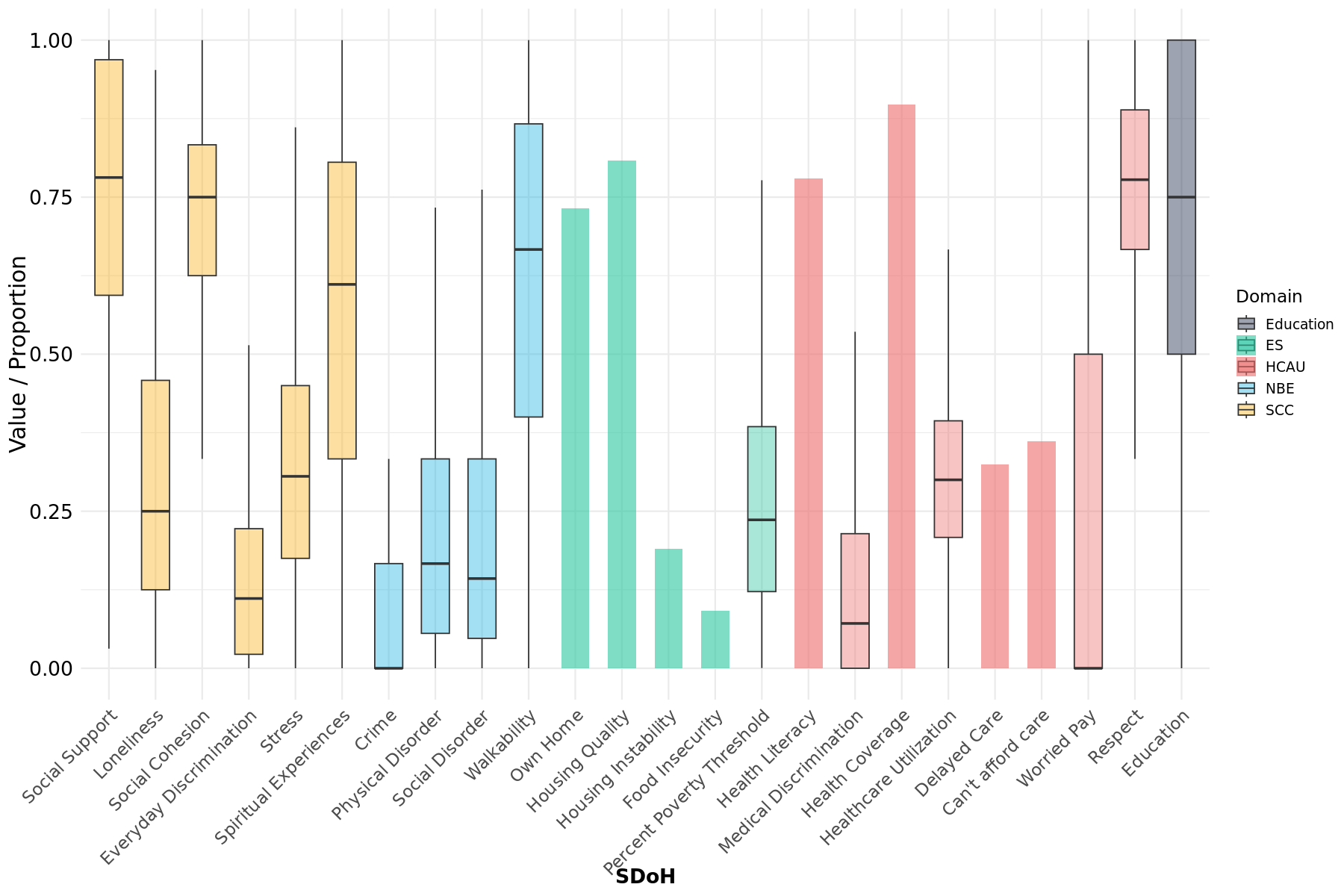

**Figure 2: Distribution of Individual-level SDoH**

Box plots depicting the distribution of individual-level SDoH in the Individual Cohort. Binary variables are represented as bar chart proportions. SDoH are color coded by HP2030 domain.

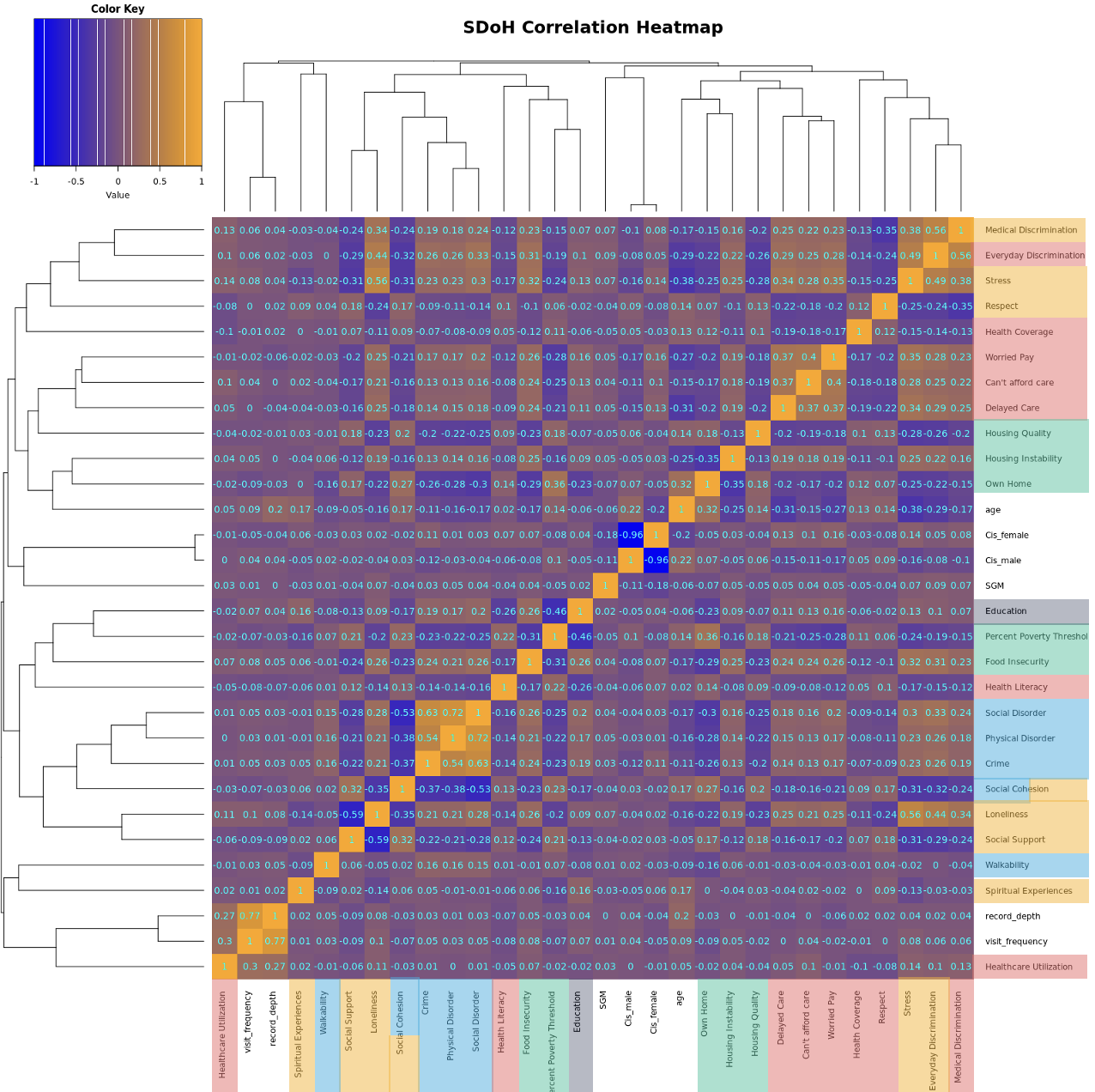
**Figure 3: Correlation Heatmap of Social Determinants of Health and Covariates Included in Study**

Each cell represents the Pearson correlation coefficient between two domain-level composite variables derived from the All of Us survey data. Orange indicates positive correlations, while blue indicates negative correlations. Items are colored by HP2030 domain.

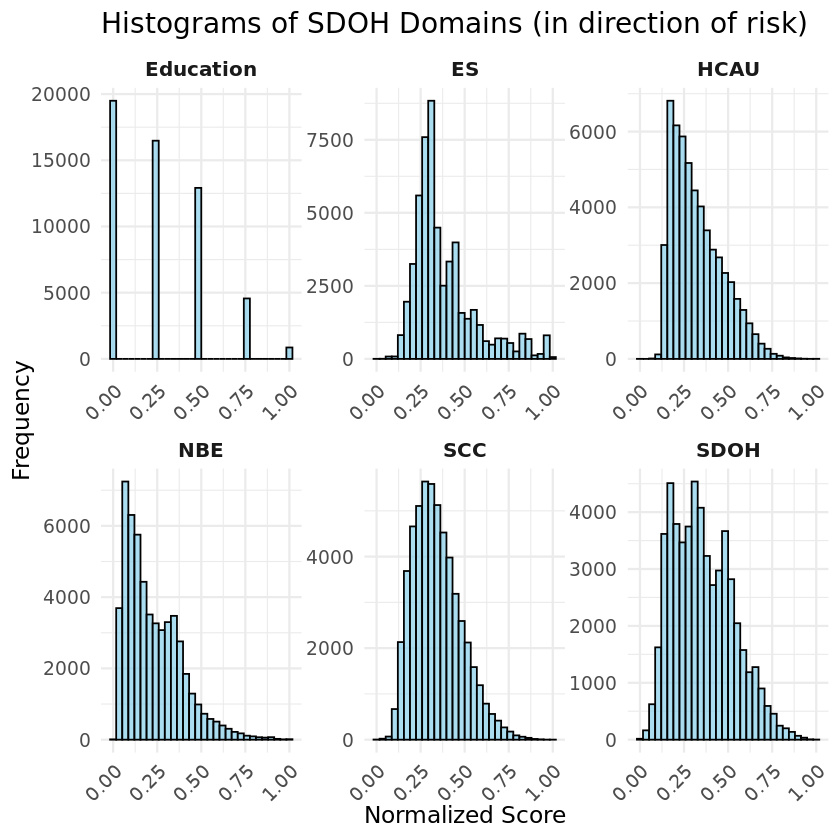

**Figure 4: Histograms of SDoH Domains**

Normalized domain scores derived from CFA for each latent SDoH domain, as well as an overall SDoH score. Scores have been rescaled to a 0–1 range for direct comparison across domains and directionally aligned so that higher values consistently indicate greater social disadvantage or risk. For interpretability, this means that, for example, a higher HCAU score reflects lower access to healthcare. The NBE domain did not require flipping. For the education domain, a score of 1 corresponds to having less than a high school education, while a score of 0 reflects a college degree or higher.

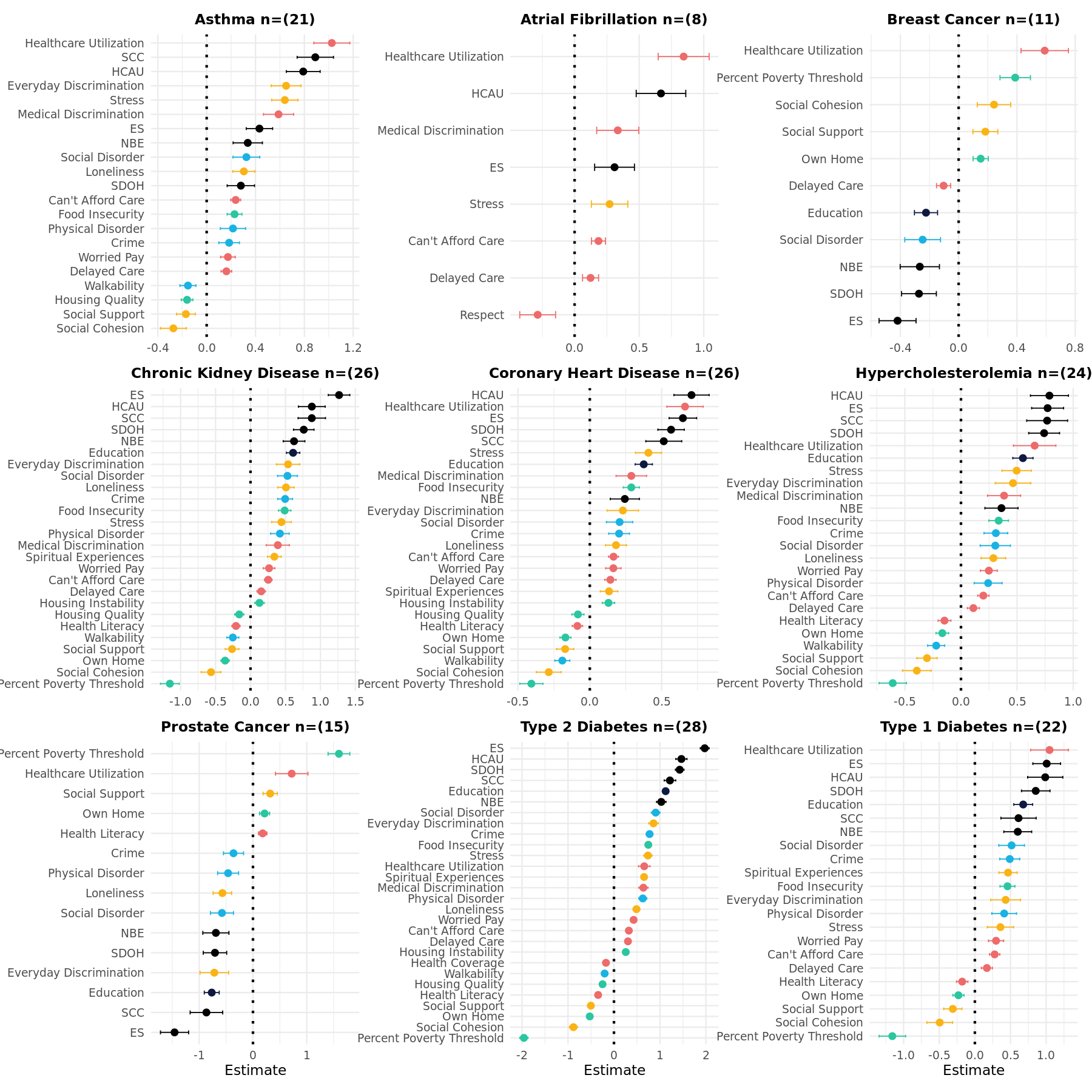

**Figure 5: Significant Associations Between Individual-level SDoH and Disease Outcomes**

Each panel displays the estimate and standard error for each SDoH significantly associated with the disease (Bonferroni corrected p-value < (.05/(29 tested variables*9 disease outcomes))). The number of significant associations is in parenthesis in the title of each disease. Each panel is ordered independently by strength of association. Domains are colored by type in accordance with the HP2030 coloring in Figure 1, and with black indicating domain-level factor scores derived from CFA. Domains are in the direction of risk. Imputed individual component predictors were used to maintain a consistent sample. Within each disease, domains are ordered from highest to lowest estimated effect size. Points indicate the magnitude and direction of association; horizontal lines represent 95% confidence intervals. The dotted vertical line at zero marks the null effect.

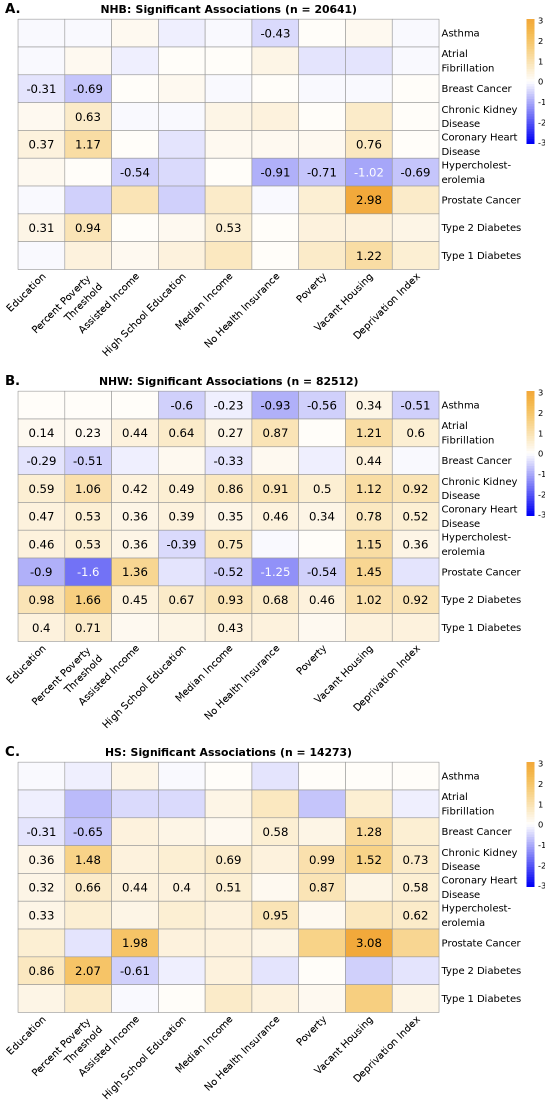

**Figure 6: Heatmaps of Significant Associations by SIRE**

Heatmap displaying the estimates from SEMs assessing the relationship between SDoH metrics and nine chronic conditions in the SES Cohort in the NHB population **(A)**, the NHW population **(B)**, and the HS population **(C)**. Only statistically significant associations are annotated with effect sizes. Negative values indicate that higher levels in a specific SDoH risk are associated with reduced probability of disease. Heatmaps are on the same scale to facilitate direct comparison between individual-level and area-level SDoH associations.

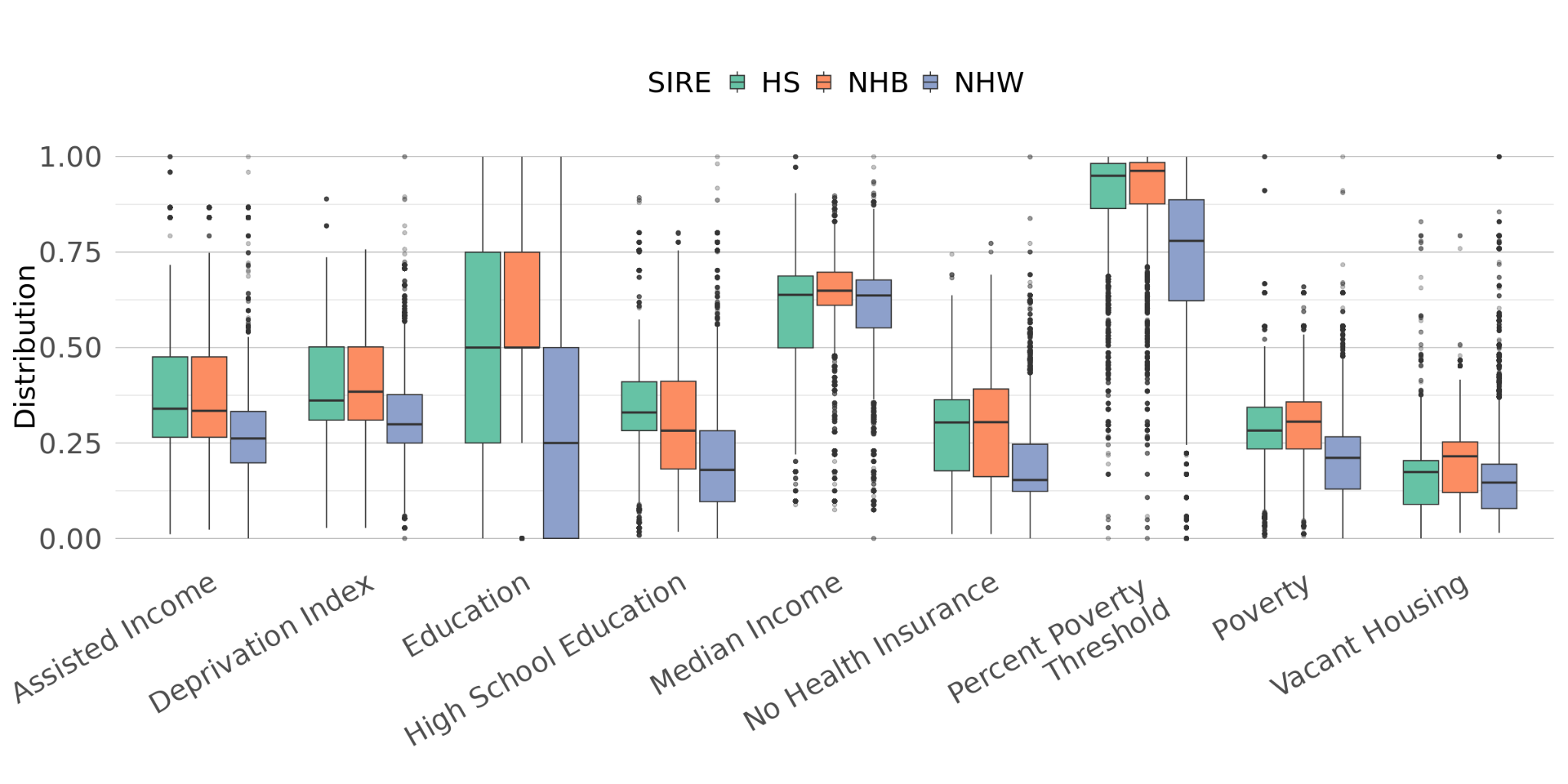

**Figure 7: Distributions of SDoH in SES-SIRE Cohort across SIRE**

Box plots of SDoH metrics in the SES-SIRE Cohort stratified by SIRE. Boxes are color-coded by SIRE. All values were min-max normalized to a scale of 0-1 and flipped into the direction of increased risk to facilitate direct comparison.

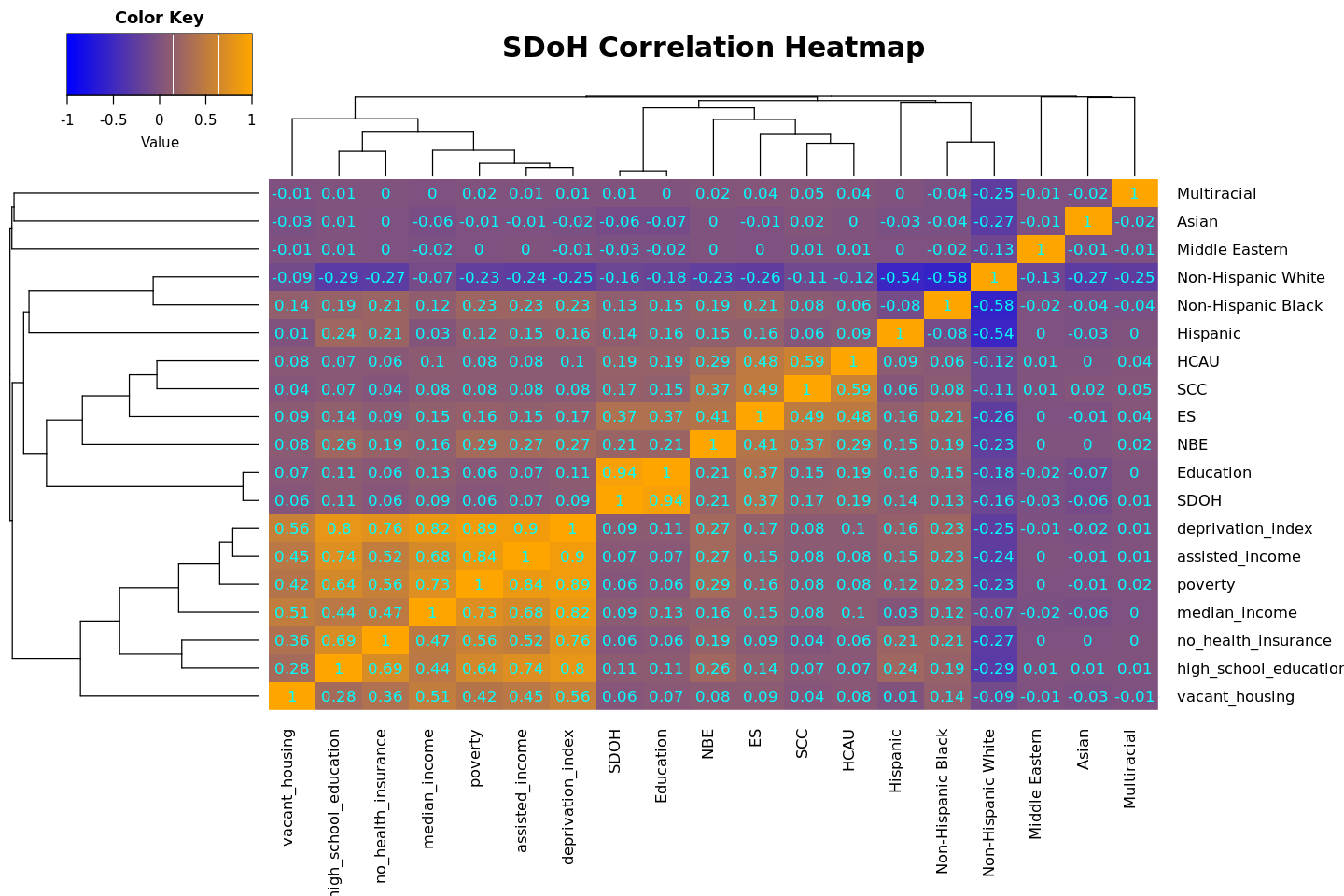

**Figure 8: SDoH Correlation Heatmap with SIRE**

Correlation heatmap of individual- and areal-level SDoH with SIRE. Each cell represents the Pearson correlation coefficient. Orange indicates positive correlations, while blue indicates negative correlations.

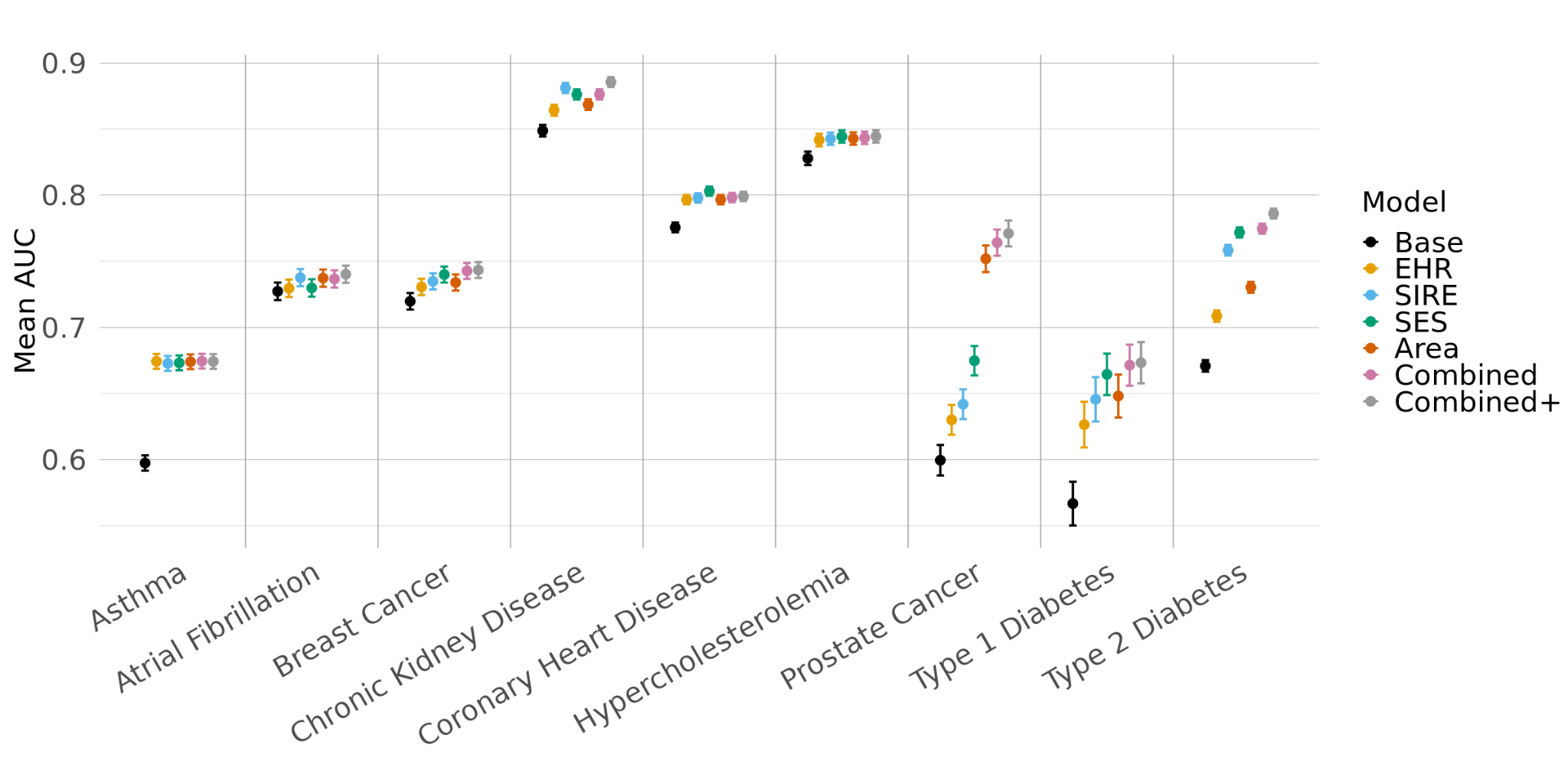
 **Figure 9: Mean AUC of SDoH and non-SDoH Elastic Net Models Across Nine Chronic Conditions in SES Cohort**

Dot plot showing the mean AUC and corresponding 95% confidence intervals from 5-fold cross-validated elastic net models using a 70/30 training/testing split. The x-axis lists the nine chronic conditions, while each point is color coded by a different model type. The Base model includes age, age2, and Sex/Gender. In addition to the covariates included in the Base model, the EHR model also includes visit frequency and record depth. The rest of the models build on the EHR mode. The SIRE model also includes NHB and HS. The SES model also includes percent of poverty threshold and education. The Area model includes the PsRS trained on all area-level metrics. The Combined model includes a PsRS trained on the percent of poverty threshold, education, and area-level metrics and the Combined+ model additionally includes NHB and HS.

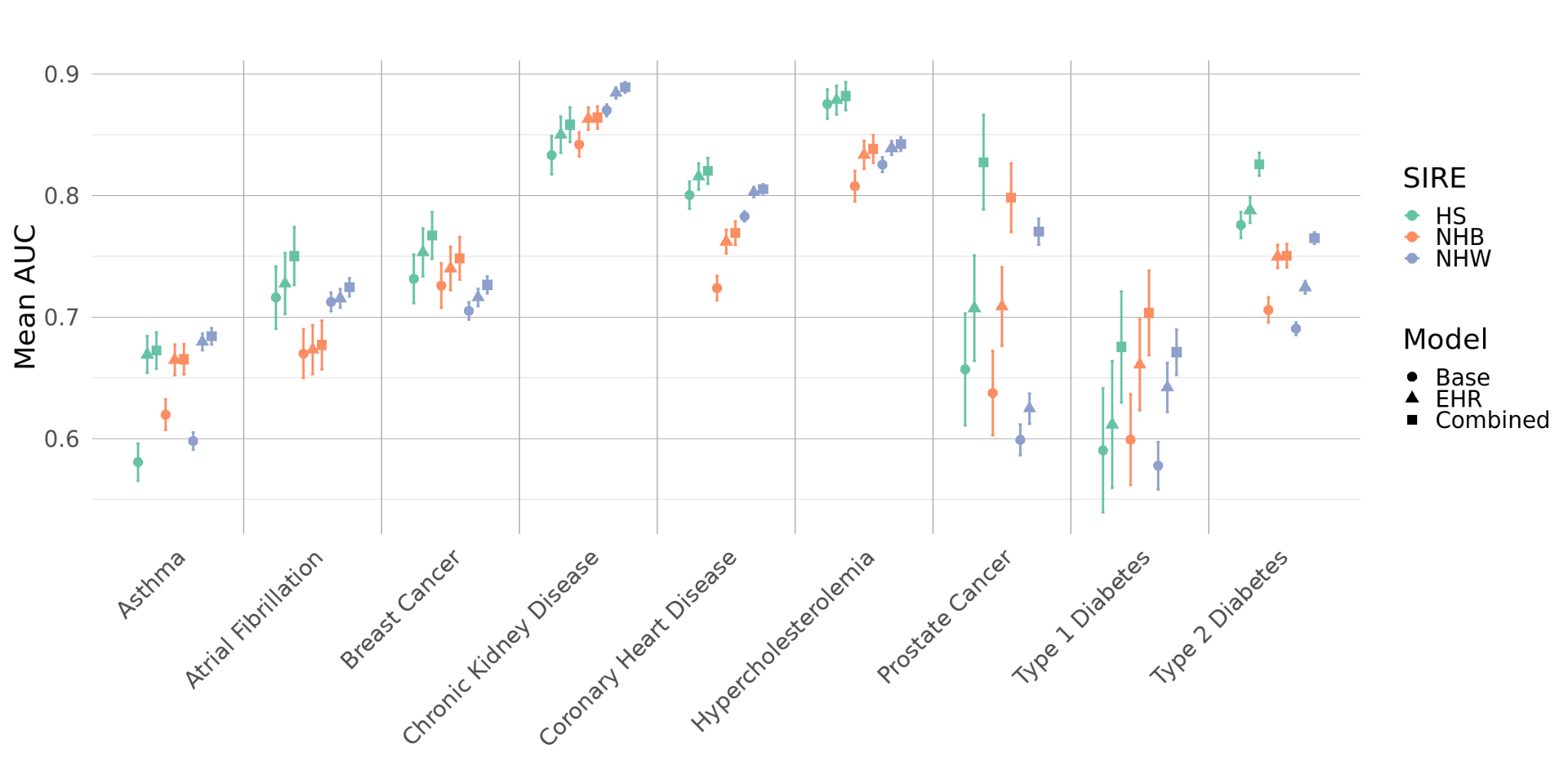

**Figure 10: Comparison of model performance across SIRE for Nine Chronic in SES Cohort**

Dot plot showing the mean AUC and corresponding 95% confidence intervals from 5-fold cross-validated elastic net models using a 70/30 training/testing split. The x-axis lists the nine chronic conditions, while each point represents a different model type and SIRE. Models are color coded by SIRE and shapes are used to delineate model types.

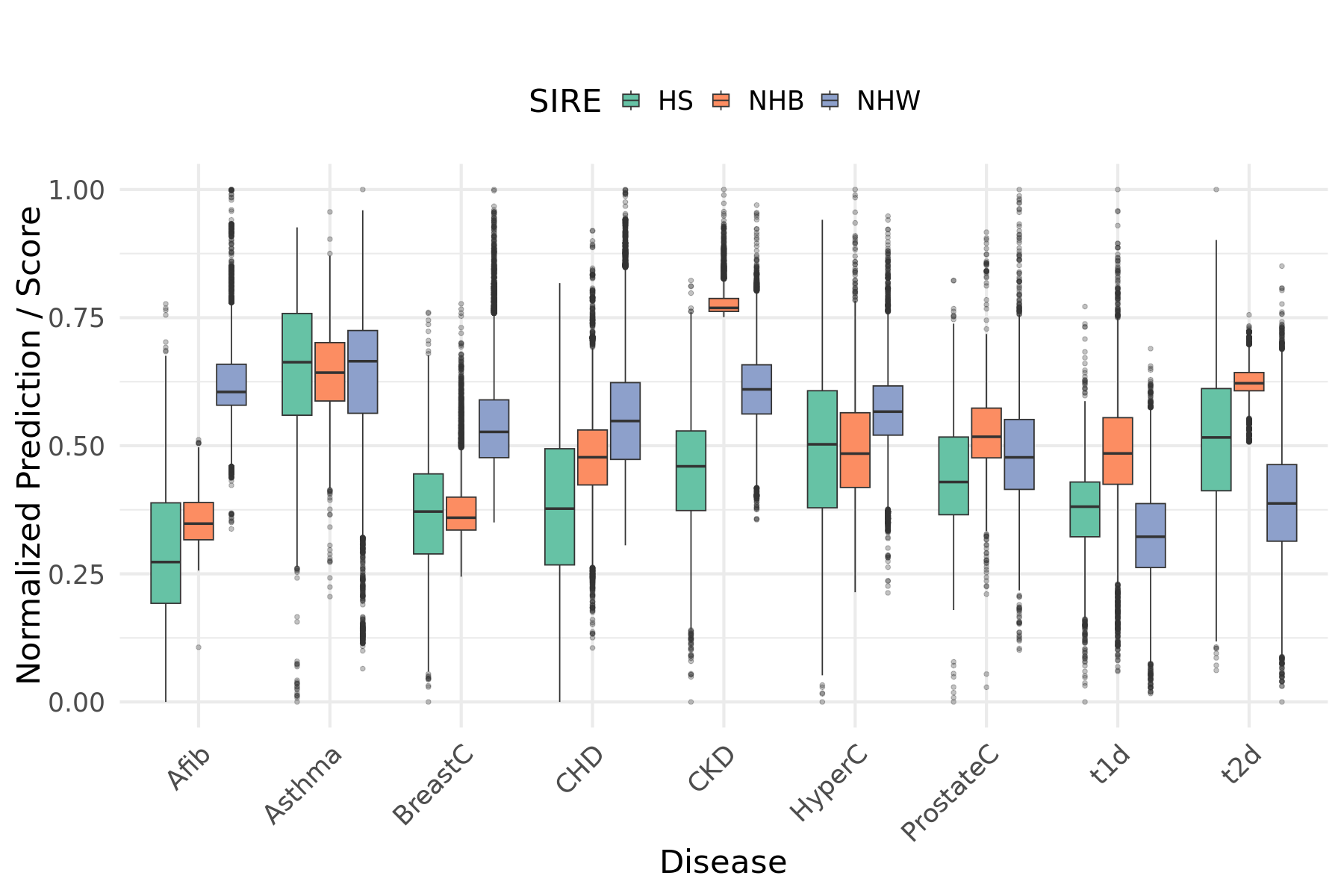

**Figure 11: Normalized distributions of stratified Combined PsRS across diseases**

Box plots of Combined PsRS stratified by SIRE across diseases. Combined PsRS were calculated independently in stratified analyses and then were min-max normalized to a 0-1 scale by disease across all populations. Boxes are colored by SIRE.

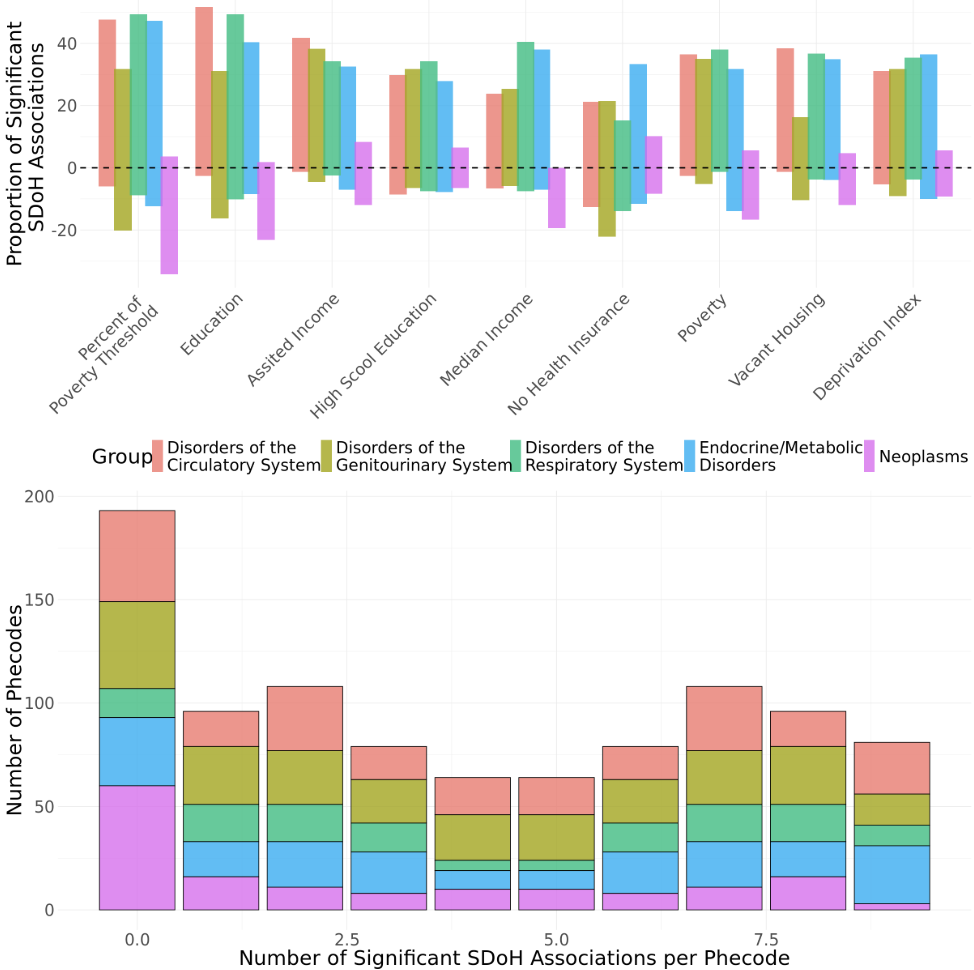

**Figure 12: Targeted Phenome-Wide Association Study of SDOH in SES Cohort**

**A**. Box plot showing the proportion of significant SDoH associations for each disease group. The y-axis represents the proportion of significant associations, split into positive (above the x-axis, associated with increased disease prevalence) and negative (below the x-axis, associated with decreased disease prevalence) effects. Boxes are grouped by SDoH category and color-coded by disease group. **B.** Histogram showing the distribution of significant SDoH associations across phecodes. The x-axis indicates the number of SDoH significantly associated with each phecode (maximum SDoH tested = 9), while the y-axis shows the count of phecodes with that number of associations.
