## Appendix I for "Quantifying Social Determinants of Health for Disease Prediction: A Multi-Level Approach Using Healthy People 2030 and All of Us Data"

**Economic Stability	2**

Percent Poverty Threshold	2

Food Insecurity	2

Housing Instability	3

Home Ownership	3

Housing Quality	4

**Education Access and Quality	4**

Educational Attainment	4

**Health Care Access and Utilization	5**

Medical Discrimination	5

Health Literacy	5

Health Care Coverage	6

Health Care Utilization	6

Delay Care	7

Can’t Afford Care	7

Worried Paying Medical Bills	8

Respected by Provider	8

**Neighborhood and Built Environment	9**

Neighborhood Physical Disorder	9

Neighborhood Social Disorder	9

Neighborhood Walkability	10

Neighborhood Crime	10

**Social and Community Context	11**

Social Cohesion Among Neighbors	11

Social Support	11

Loneliness	12

Everyday Discrimination	12

Stress	12

Spiritual Experiences	13

### Economic Stability

#### Percent Poverty Threshold

**Original source:** Office of the Assistant Secretary for Planning and Evaluation 2017 Poverty Guidelines: <https://aspe.hhs.gov/topics/poverty-economic-mobility/poverty-guidelines/prior-hhs-poverty-guidelines-federal-register-references/2017-poverty-guidelines#threshholds> (<https://aspe.hhs.gov/sites/default/files/documents/3edbd42a9b8de4f2a87211283e541ca4/historical-poverty-guidelines-through-2024.xlsx>)

**All of Us source:** [The Basics Survey](https://www.researchallofus.org/wp-content/themes/research-hub-wordpress-theme/media/surveys/Survey_Basics_Eng_Src.pdf)

**Items: “**What is your annual household income from all sources?”

**Options:** Less than $10,000, $10,000- $24,999, $25,000-$34,999, $35,000-$49,999, $50,000- $74,999, $75,000-$99,999, $100,000- $149,999, $150,000- $199,999, $200,000 or more, Prefer not to answer

**Scoring:** Less than $10,000 = 5,000, $10,000- $24,999 = 17,500k, $25,000-$34,999 = 30,000, $35,000-$49,999 = 42,500, $50,000- $74,999 = 62,500, $75,000-$99,999 = 87,500, $100,000- $149,999 = 125,000, $150,000- $199,999 = 175,000, $200,000 or more = 225,000. These integer values were then divided by the corresponding poverty threshold based on location (48 continuous states, Hawaii, or Alaska), size of household, and month/year they filled out the survey. The resulting value was then multiplied by 100 to get a percentage of the poverty threshold. The nonfarm data was used since we had no way of knowing which participants were registered as owning a farm.

**Code shorthand:** per_poverty_threshold

#### Food Insecurity

**Original source:** [Hager et al., 2012](https://publications.aap.org/pediatrics/article-abstract/126/1/e26/68243/Development-and-Validity-of-a-2-Item-Screen-to?redirectedFrom=fulltext); [Radandt et al., 2018](https://www.ncbi.nlm.nih.gov/pmc/articles/PMC6419517/) (2-item)

**All of Us source:** [Social Determinants of Health Survey](https://www.researchallofus.org/wp-content/themes/research-hub-wordpress-theme/media/surveys/Survey_SDOH_Eng_Src.pdf)

**Items:**

- “Within the past 12 months, we worried whether our food would run out before we got money to buy more.”
- “Within the past 12 months, the food we bought just didn’t last and we didn’t have money to get more.”

**Options:** 3-point Options (often true, sometimes true or never true)

**Scoring:** An individual is considered food insecure if any item is rated as sometimes true or often true. Food insecure was assigned a value of 1.

**Cronbach's alpha (SDoH cohort):** 0.79 [0.790, 0.797]

**Cronbach's alpha (genetics cohort):** 0.79 [0.782, 0.790]

**Code shorthand:** food_insecurity

#### Housing Instability

**Original source:** Manchanda, R. & Gottlieb, L. Upstream Risks Screening Tool and Guide V2.6. https://www.aamc.org/media/25736/download (2015). (1-item)

**All of Us source:** [Social Determinants of Health Survey](https://www.researchallofus.org/wp-content/themes/research-hub-wordpress-theme/media/surveys/Survey_SDOH_Eng_Src.pdf)

**Items: “**In the last 12 months, how many times have you or your family moved from one home to another?”

**Options:** Fill in the blank integer value

**Scoring:** Binary: 1 for 0 moves or 0 for 1 or more moves

**Original source:** UK Biobank. Stockport, UK: UK Biobank. Available from: <https://www.ukbiobank.ac.uk/> 2011

**All of Us source:** [The Basics Survey](https://www.researchallofus.org/wp-content/themes/research-hub-wordpress-theme/media/surveys/Survey_Basics_Eng_Src.pdf)

**Items: “**How many years have you lived at your current address?”

**Options:** Less than 1 year, 1-2 years, 3-5 years, 6-10 years, 11-20 years, More than 20 years

**Scoring:** Binary: 1 or 0 where 1 is less than a year and all other options = 0

**Original source:** The Homeless Screener Clinical Reminder (HSCR). Washington DC: U.S. Department of Veteran Affairs. Available from:

<https://www.va.gov/HOMELESS/nchav/resources/prevention/homeless-screener.asp> 2012

**All of Us source:** [The Basics Survey](https://www.researchallofus.org/wp-content/themes/research-hub-wordpress-theme/media/surveys/Survey_Basics_Eng_Src.pdf)

**Items: “**In the past 6 months, have you been worried or concerned about NOT having a place to live?”

**Options:** Yes, No

**Scoring:** Yes = 1, No = 0

**Section Scoring:** 0 = housing insecurity if anyone had a 0 for any category

**Cronbach's alpha (SDoH cohort):** 0.79 [0.784, 0.790]

**Cronbach's alpha (genetics cohort):** 0.79 [0.783, 0.790]

**Code shorthand:** housing_instability

#### Home Ownership

**Original source:** Manchanda, R. & Gottlieb, L. Upstream Risks Screening Tool and Guide V2.6. https://www.aamc.org/media/25736/download (2015). (1-item)

**All of Us source:** [Social Determinants of Health Survey](https://www.researchallofus.org/wp-content/themes/research-hub-wordpress-theme/media/surveys/Survey_SDOH_Eng_Src.pdf)

**Items: “**Do you own or rent the place where you live?”

**Options:** Own, Rent, Other arrangement, Prefer not to answer. Branching logic: when “Other arrangement” selected, then: On a college campus, With a friend/roommate, With family, Motel/hotel, Hospital, rehabilitation center, drug treatment center, or other temporary institution, In a group home, nursing home, or other residential facility, Transitional housing, Emergency shelter or homeless shelter, Anywhere outside (e.g., street, vehicle, abandoned building), Other

**Scoring:** Binary 0 or 1 where 1=Home Own and 0=anything else

**Code shorthand:** home_own

#### Housing Quality

**Original source:** [Billioux et al., 2017](https://nam.edu/standardized-screening-for-health-related-social-needs-in-clinical-settings-the-accountable-health-communities-screening-tool/)

**All of Us source:** [Social Determinants of Health Survey](https://www.researchallofus.org/wp-content/themes/research-hub-wordpress-theme/media/surveys/Survey_SDOH_Eng_Src.pdf)

**Items: “**Think about the place you live. Do you have problems with any of the following?”

**Options:** Check all that apply: Bug infestation, Mold, Lead paint or pipes, Inadequate heat, Oven or stove not working, No or not working smoke detector, Water leaks, None of the above

**Scoring:** Binary 0 or 1 or more where 1 indicates good housing quality

**Code shorthand:** housing_quality

### Education Access and Quality

#### Educational Attainment

**Original source:** Behavioral Risk Factor Surveillance System (BRFSS). Atlanta, GA: Centers for Disease Control and Prevention. Available from:

<https://www.cdc.gov/brfss/questionnaires/index.htm> 2016

**All of Us source:** [The Basics Survey](https://www.researchallofus.org/wp-content/themes/research-hub-wordpress-theme/media/surveys/Survey_Basics_Eng_Src.pdf)

**Items: “**What is the highest grade or year of school you completed?”

**Options:** Never attended school or only attended kindergarten, Grades 1 through 4 (Primary), Grades 5 through 8 (Middle school), Grades 9 through 11 (Some high school), Grade 12 or GED (High school graduate), 1 to 3 years after high school (Some college, Associate’s degree, or technical school), College 4 years or more (College graduate), Advanced degree (Master’s, Doctorate, etc.), Prefer not to answer

**Scoring:** Less than high school = 1, High school graduate = 2, 1 to 3 years after high school = 3, 4 or more years college = 4, Advance degree = 5

**Code shorthand:** education

### Health Care Access and Utilization

#### Medical Discrimination

**Original source:** Everyday discrimination Options in medical settings (7-item) [Peek et al., 2011](https://www.ncbi.nlm.nih.gov/pmc/articles/PMC3350778/)

**All of Us source:** [Social Determinants of Health Survey](https://www.researchallofus.org/wp-content/themes/research-hub-wordpress-theme/media/surveys/Survey_SDOH_Eng_Src.pdf)

**Items:**

- “You are treated with less courtesy than other people are.”
- “You are treated with less respect than other people are.”
- “You receive poorer service than others.”
- “A doctor or nurse acts if he or she thinks you are not smart.”
- “A doctor or nurse acts as if he or she is afraid of you.”
- “A doctor or nurse acts as if he or she is better than you.”
- “You feel like a doctor or nurse is not listening to what you were saying.”

**Options:** 5-point Options (1 = Never; 2 = Rarely; 3 = Sometimes; 4 = Most of the time; 5 = Always)

**Scoring:** average, and the Options is flagged as missing if more than two items do not have valid responses.

**Cronbach's alpha (SDoH cohort):** 0.90 [0.894, 0.897]

**Cronbach's alpha (genetics cohort):** 0.90 [0.894, 0.897]

**Code shorthand:** health_discrim

#### Health Literacy

**Original source:** [Chew, Bradley, & Boyko, 2004](https://pubmed.ncbi.nlm.nih.gov/15343421/)

**All of Us source:** [Overall Health Survey](https://www.researchallofus.org/wp-content/themes/research-hub-wordpress-theme/media/surveys/Survey_OverallHealth_Eng_Src.pdf)

**Items:**

- “How confident are you filling out medical forms by yourself?”
- “How often do you have someone help you read health-related materials?”
- “How often do you have problems learning about your medical condition because of

difficulty understanding written information?”

**Options:** 5-point scale (0 = Not at all; 1 = A little bit; 2 = Somewhat; 3 = Quite a bit; 4 = Extremely) (0 = Always, 1 = Often, 2 = Sometimes, 3 = Occasionally, 4 = Never)

**Scoring:** mean of the three items - flagged as missing if one of the three items was missing a valid response. Health Literacy had a skewness of -2.61 and a kurtosis of 9.01, indicating it was not normal and failed our model assumptions. To remedy this, it was converted to a binary indicator variable. Individuals with a score greater than 3.5 were said to have high health literacy whereas individuals with a score lower than 3.5 were said to have low health literacy. High health literacy = 1; low health literacy = 0.

**Cronbach's alpha (SDoH cohort):** 0.69 [.688, 0.697]

**Cronbach's alpha (genetics cohort):** 0.69 [0.684, 0.693]

**Code shorthand:** health_literacy

#### Health Care Coverage

**Original source:** National Health and Nutrition Examination Survey (NHANES). Hyattsville, MD: National Center for Health Statistics. Available from: <https://wwwn.cdc.gov/nchs/nhanes/continuousnhanes/questionnaires.aspx?BeginYear=2017>

**All of Us source:** [The Basics Survey](https://www.researchallofus.org/wp-content/themes/research-hub-wordpress-theme/media/surveys/Survey_Basics_Eng_Src.pdf)

**Items: “**Are you covered by health insurance or some other kind of health care plan?”

**Options:** Yes, No. If yes, branching question: Are you currently covered by any of the following types of health insurance or health coverage plans? Select all that apply from one group

**Scoring:** Yes =1, No=0

**Original source:** National Health Interview Survey (NHIS). Hyattsville, MD: National Center for Health Statistics. Available from: <https://www.cdc.gov/nchs/nhis/nhis_questionnaires.htm> 2016

**All of Us source:** [Health Care Access & Utilization Survey](https://www.researchallofus.org/wp-content/themes/research-hub-wordpress-theme/media/surveys/Survey_HealthCare_Eng_Src.pdf)

**Items: “**DURING THE PAST 12 MONTHS, were you told by a health care provider or doctor’s office that they did not accept your health care coverage?”

**Options:** Yes, No

**Scoring:** Yes = 0, No = 1

**Section scoring:** If 0 for either question, 0 given to score. If 1 for either option, 1 given to score

**Code shorthand:** health_coverage

#### Health Care Utilization

**Original source:** National Health Interview Survey (NHIS). Hyattsville, MD: National Center for Health Statistics. Available from: <https://www.cdc.gov/nchs/nhis/nhis_questionnaires.htm> 2016

**All of Us source:** [Health Care Access & Utilization Survey](https://www.researchallofus.org/wp-content/themes/research-hub-wordpress-theme/media/surveys/Survey_HealthCare_Eng_Src.pdf)

**Items: “**DURING THE PAST 12 MONTHS, have you seen or talked to any of the following doctors or health care providers about your own health”:

- “General doctor: A provider who sees adult patients for wellness exams and the treatment of diseases.”
- “A nurse practitioner, physician assistant, or midwife?”
- “A doctor who specializes in women's health (an obstetrician/gynecologist)?”
- “A mental health professional such as a psychiatrist, psychologist, psychiatric nurse, or clinical social worker?”
- “An optometrist, ophthalmologist, or eye doctor (someone who prescribes eyeglasses)?”
- “A podiatrist or foot doctor?”
- “A chiropractor?”
- “A physical therapist, speech therapist, respiratory therapist, audiologist, or occupational therapist?”
- “A dentist or orthodontist?”
- “A medical doctor who specializes in a particular medical disease or problem (other than obstetrician/gynecologist, psychiatrist, or ophthalmologist)?”
- “A specialist: A provider who treats patients for diseases related to a particular system of the body, such as a cardiologist or urologist”
- “Traditional healers such as, Shaman, acupuncturist, or non-western medicine?”

**Options:** None, 1, 2 to 3, 4 to 5, 6 to 7, 8 to 9, 10 to 12, 13 to 15, 16 or More

**Scoring:** 0 = None, 1 = 1, 2 = 2 to 3, 3 = 4+; items averaged - The Options is set to missing if more than four items did not have a valid response

**Cronbach's alpha (SDoH cohort):** 0.54 [0.532, 0.545]

**Cronbach's alpha (genetics cohort):** 0.54 [0.530, 0.544]

**Code shorthand:** health_care_use

#### Delay Care

**Original source:** National Health Interview Survey (NHIS). Hyattsville, MD: National Center for Health Statistics. Available from: <https://www.cdc.gov/nchs/nhis/nhis_questionnaires.htm> 2016

**All of Us source:** [Health Care Access & Utilization Survey](https://www.researchallofus.org/wp-content/themes/research-hub-wordpress-theme/media/surveys/Survey_HealthCare_Eng_Src.pdf)

**Items: “**There are many reasons people delay getting medical care. Have you delayed getting care for any of the following reasons in the PAST 12 MONTHS?”

- “Couldn’t get child care.”*
- “You provide care to an adult and could not leave him/her.”*
- “You live in a rural area where distance to the health care provider is too far”*
- “Didn’t have transportation”*
- “Couldn’t get time off work.”
- “You were nervous about seeing a health care provider.”
- “Couldn’t afford the copay.”
- “Your deductible was too high/or could not afford the deductible.”
- “You had to pay out of pocket for some or all of the procedure.”

**Options:** Yes, No, don’t know

**Scoring:** if at least 7 options filled in 1 given if answered Yes to any question

*converted to binary to match phenxtoolkit

**Original Cronbach's alpha (SDoH cohort):** 0.62 [0.616 0.626]

**Original Cronbach's alpha (genetics cohort):** 0.62 [0.615 0.626]

*To improve interscale reliability, removed items with Item-Total Correlation below 0.3 which were: “Couldn’t get child care.” “You provide care to an adult and could not leave him/her.” “You live in a rural area where distance to the health care provider is too far” “Didn’t have transportation”

**Cronbach's alpha (SDoH cohort):** 0.76 [0.751, 0.758]

**Updated Cronbach's alpha (genetics cohort):** 0.76 [ [0.752, 0.759]

**Code shorthand:** delayed_care

#### Can’t Afford Care

**Original source:** National Health Interview Survey (NHIS). Hyattsville, MD: National Center for Health Statistics. Available from: <https://www.cdc.gov/nchs/nhis/nhis_questionnaires.htm> 2016

**All of Us source:** [Health Care Access & Utilization Survey](https://www.researchallofus.org/wp-content/themes/research-hub-wordpress-theme/media/surveys/Survey_HealthCare_Eng_Src.pdf)

**Items:** “DURING THE PAST 12 MONTHS, was there any time when you needed any of the following, but didn't get it because you couldn't afford it?”

- “Prescription medicines”
- “Mental health care or counseling”
- “Emergency care”
- “Dental care (including check ups)”
- “Eyeglasses”
- “To see a regular doctor or general health provider (in primary care, general practice, internal medicine, family medicine)”
- “To see a specialist”
- “Follow-up care”

“DURING THE PAST 12 MONTHS, were any of the following true for you?”

- “You skipped medication doses to save money”
- “You took less medicine to save money” “
- You delayed filling a prescription to save money”
- “You asked your doctor for a lower cost medication to save money”
- “You bought prescription drugs from another country to save money”
- “You used alternative therapies to save money”

**Options:** Yes, No, Don’t know

**Scoring:** if at least 11 options filled in 0 given if answered Yes to any question

*converted to binary to match phenxtoolkit

**Cronbach's alpha (genetics cohort):** 0.84 [0.836, 0.841]

**Cronbach's alpha (SDoH cohort):** 0.84 [0.835, 0.840]

**Code shorthand:** afford_care

#### Worried Paying Medical Bills

**Original source:** National Health Interview Survey (NHIS). Hyattsville, MD: National Center for Health Statistics. Available from: <https://www.cdc.gov/nchs/nhis/nhis_questionnaires.htm> 2016

**All of Us source:** [Health Care Access & Utilization Survey](https://www.researchallofus.org/wp-content/themes/research-hub-wordpress-theme/media/surveys/Survey_HealthCare_Eng_Src.pdf)

**Items:** “If you get sick or have an accident, how worried are you that you will be able to pay your medical bills? Are you very worried, somewhat worried, or not at all worried?”

**Options:** very worried, somewhat worried, or not at all worried

**Scoring:** 2 = very worried, 1 = somewhat worried, 0 = or not at all worried

**Code shorthand:** worried_pay

#### Respected by Provider

**Original source:** National Health Interview Survey (NHIS). Hyattsville, MD: National Center for Health Statistics. Available from: <https://www.cdc.gov/nchs/nhis/nhis_questionnaires.htm> 2016

**All of Us source:** [Health Care Access & Utilization Survey](https://www.researchallofus.org/wp-content/themes/research-hub-wordpress-theme/media/surveys/Survey_HealthCare_Eng_Src.pdf)

**Items:**

- “How often were you treated with respect by your doctors or health care providers? Would you say…”
- “How often did your doctors or health care providers ask for your opinions or beliefs about your medical care or treatment? For example, what kind of tests, procedures, or medications you prefer. Would you say…”
- “How often did your doctors or health care providers tell or give you information about your health and health care that was easy to understand? Would you say…”

**Options:** Always, Most of the time, some of the time, none of the time

**Scoring:** Always = 2, most of the time = 1, some or none of the time = 0; average of 3 items - The Options is set to missing if more than one item does not have a valid response

**Cronbach's alpha (SDoH cohort):** 0.60 [0.593, 0.605]

**Cronbach's alpha (genetics cohort):** 0.60 [0.590, 0.600]

**Code shorthand:** respect

### Neighborhood and Built Environment

#### Neighborhood Physical Disorder

**Original source:** Ross–Mirowsky perceived neighborhood disorder Options (6-item physical disorder subOptions) [Ross and Mirowsky, 2001](https://pubmed.ncbi.nlm.nih.gov/11668773/)

**All of Us source:** [Social Determinants of Health Survey](https://www.researchallofus.org/wp-content/themes/research-hub-wordpress-theme/media/surveys/Survey_SDOH_Eng_Src.pdf)

**Items:**

- “There is a lot of graffiti in my neighborhood.”
- “My neighborhood is noisy.”
- “Vandalism is common in my neighborhood.”
- “There are lot of abandoned buildings in my neighborhood.”
- “My neighborhood is clean.”
- “People in my neighborhood take good care of their houses and apartments.”

**Options:** 4-point agreement Options (1 = strongly disagree; 2 = disagree; 3 = agree; 4 = strongly agree)

**Scoring:** Average of 6 items with 2 reverse coded - The Options is set to missing if more than one item does not have a valid response

**Cronbach's alpha (SDoH cohort):** 0.84 [0.834, 0.838]

**Cronbach's alpha (genetics cohort):** 0.84 [0.833, 0.837]

**Code shorthand:** npd_scale

#### Neighborhood Social Disorder

**Original source:** Ross–Mirowsky perceived neighborhood disorder Options (7-item social disorder subOptions) [Ross and Mirowsky, 2001](https://pubmed.ncbi.nlm.nih.gov/11668773/)

**All of Us source:** [Social Determinants of Health Survey](https://www.researchallofus.org/wp-content/themes/research-hub-wordpress-theme/media/surveys/Survey_SDOH_Eng_Src.pdf)

**Items:**

- “There are too many people hanging around on the streets near my home.”
- “There is a lot of crime in my neighborhood.”
- “There is too much drug use in my neighborhood.”
- “There is too much alcohol use in my neighborhood.”
- “I’m always having trouble with my neighbors.”
- “In my neighborhood, people watch out for each other.”
- “My neighborhood is safe.”

**Options:** 4-point agreement rating Options (1 = strongly disagree; 2 = disagree; 3 = agree; 4 = strongly agree)

**Scoring:** Average of 7 items with 2 reverse coded - The Options is set to missing if more than one item does not have a valid response

**Cronbach's alpha (SDoH cohort):** 0.87 [0.869, 0.873]

**Cronbach's alpha (genetics cohort):** 0.87 [0.869, 0.873]

**Code shorthand:** nsd_scale

#### Neighborhood Walkability

**Original source:** Physical activity neighborhood environment Options (PANES) (5 core items) [Sallis et al., 2009](https://pubmed.ncbi.nlm.nih.gov/19460656/) [Sallis et al., 2010](https://journals.humankinetics.com/view/journals/jpah/7/4/article-p533.xml)

**All of Us source:** [Social Determinants of Health Survey](https://www.researchallofus.org/wp-content/themes/research-hub-wordpress-theme/media/surveys/Survey_SDOH_Eng_Src.pdf)

**Items:**

- “Many shops, stores, markets or other places to buy things I need are within easy walking distance of my home. Would you say that you…”
- “It is within a 10–15 minutes walk to a transit stop (such as bus, train, trolley, or tram) from my home. Would you say that you…”
- “There are sidewalks on most of the streets in my neighborhood. Would you say that you…”
- “There are facilities to bicycle in or near my neighborhood, such as special lanes, separate paths or trails, shared use paths for cycles and pedestrians. Would you say that you…”
- “My neighborhood has several free or low-cost recreation facilities, such as parks, walking trails, bike paths, recreation centers, playgrounds, public swimming pools, etc. Would you say that you…”

**Options:** 4-point agreement Options: 1 = Strongly disagree, Does not apply to my neighborhood; 2 = Somewhat disagree; 3 = Somewhat agree; 4 = Strongly agree; Missing= Skip/Don’t know or Does not apply to my neighborhood

**Scoring:** average of the five items - flagged the score as missing if more than one item does not have a valid response; Does notes apply to my neighborhood scored as 1 = strongly disagree

**Cronbach's alpha (SDoH cohort):** 0.78 [0.770, 0.783]

**Cronbach's alpha (genetics cohort):** 0.78 [0.777, 0.783]

**Code shorthand:** Walkability

#### Neighborhood Crime

**Original source:** Physical activity neighborhood environment Options (PANES) crime(2-item subOptions) [Sallis et al., 2009](https://pubmed.ncbi.nlm.nih.gov/19460656/) [Sallis et al., 2010](https://journals.humankinetics.com/view/journals/jpah/7/4/article-p533.xml)

**All of Us source:** [Social Determinants of Health Survey](https://www.researchallofus.org/wp-content/themes/research-hub-wordpress-theme/media/surveys/Survey_SDOH_Eng_Src.pdf)

**Items:**

- “The crime rate in my neighborhood makes it unsafe to go on walks at night. Would you say that you…”
- “The crime rate in my neighborhood makes it unsafe to go on walks during the day. Would you say that you…”

**Options:** 4-point agreement Options (1= Strongly disagree; 2 = Somewhat disagree; 3 = Somewhat agree; 4 = Strongly agree; Missing= Skip/Don’t know).

**Scoring:** Averaged - either item is not a valid response (including Skip/Don’t know), we flagged the Options as missing. Additional coding strategies for individual items are recommended by Sallis et al., 2009

**Cronbach's alpha (SDoH cohort):** 0.75 [0.750, 0.759]

**Cronbach's alpha (genetics cohort):** 0.76 [0.751, 0.761]

**Code shorthand:** crime_saftey

### Social and Community Context

#### Social Cohesion Among Neighbors

**Original source:** Social cohesion neighborhood Options (4-item) [Mujahid et al., 2007](https://pubmed.ncbi.nlm.nih.gov/17329713/)

**All of Us source:** [Social Determinants of Health Survey](https://www.researchallofus.org/wp-content/themes/research-hub-wordpress-theme/media/surveys/Survey_SDOH_Eng_Src.pdf)

**Items:**

- “People around here are willing to help their neighbors.”
- “People in my neighborhood generally get along with each other.”
- “People in my neighborhood can be trusted.”
- “People in my neighborhood share the same values.”

**Options:** 5-point agreement Options (5 = strongly agree, 4 = agree, 3 = neutral (neither agree nor disagree), 2 = disagree, and 1 = strongly disagree)

**Scoring:** mean of the four items - flagged as missing if any of the four items were missing a valid response

**Cronbach's alpha (SDoH cohort):** 0.87 [0.873, 0.876]

**Cronbach's alpha (genetics cohort):** 0.87 [0.872, 0.876]

**Code shorthand:** social_cohesion

*cross-loaded with NBD in SDoH factor model

#### Social Support

**Original source:** RAND MOS social support survey (8-item) [Moser et al., 2012](https://www.ncbi.nlm.nih.gov/pmc/articles/PMC4119888/)

**All of Us source:** [Social Determinants of Health Survey](https://www.researchallofus.org/wp-content/themes/research-hub-wordpress-theme/media/surveys/Survey_SDOH_Eng_Src.pdf)

**Items:**

- “Someone to help you if you were confined to bed”
- “Someone to take you to the doctor if you need it”
- “Someone to prepare your meals if you were unable to do it yourself”
- “Someone to help with daily chores if you were sick”
- “Someone to have a good time with”
- “Someone to turn to for suggestions about how to deal with a personal problem”
- “Someone who understands your problems”
- “Someone to love and make you feel wanted”

**Options:** 5-point Options (1 = None of the time; 2 = A little of the time; 3 = Some of the time; 4 = Most of the time; 5 = All of the time)

**Scoring:** “Three scores can be computed using an average of the items

within the Options. A total social support Options can be computed using all eight items. It can be computed if 6 or more items have valid responses. There are also two 4-item subOptionss, one to measure instrumental support (e.g., help prepare your meals), and the other to measure emotional support (e.g., someone to love and make you feel wanted). These Options are computed as averages and

can be used if only a single item is missing, otherwise the Options is set to a missing value.”

**Cronbach's alpha (SDoH cohort):** 0.95 [0.948, 0.949]

**Cronbach's alpha (genetics cohort):** 0.95 [0.948, 0.950]

**Code shorthand:** social_support

#### Loneliness

**Original source:** UCLA loneliness Options (8-item) [Hays & DiMatteo et al., 1987](https://pubmed.ncbi.nlm.nih.gov/3572711/)

**All of Us source:** [Social Determinants of Health Survey](https://www.researchallofus.org/wp-content/themes/research-hub-wordpress-theme/media/surveys/Survey_SDOH_Eng_Src.pdf)

**Items:**

- “I lack companionship”
- “There is no one I can turn to”
- “I am an outgoing person”
- “I feel left out”
- “I feel isolated from others”
- “I can find companionship when I want it”
- “I am unhappy being so withdrawn”
- “People are around me but not with me”

**Options:** 4-point frequency rating (1 = Never; 2 = Rarely; 3 = Sometimes; 4 = Often)

**Scoring:** item mean and can be computed if 6 or more items have valid

Responses - rescaled to 0-100 scale

**Cronbach's alpha (SDoH cohort):** 0.87 [0.870, 0.873]

**Cronbach's alpha (genetics cohort):** 0.87 [0.870, 0.873]

**Code shorthand:** loneliness

#### Everyday Discrimination

**Original source:** Everyday discrimination Options (9-item) [Williams 1997](https://pubmed.ncbi.nlm.nih.gov/9250627/)

**All of Us source:** [Social Determinants of Health Survey](https://www.researchallofus.org/wp-content/themes/research-hub-wordpress-theme/media/surveys/Survey_SDOH_Eng_Src.pdf)

**Items:**

- “You are treated with less courtesy than other people are.”
- “You are treated with less respect than other people are.”
- “You receive poorer service than other people at restaurants or stores.”
- “People act as if they think you are not smart.”
- “People act as if they are afraid of you.”
- “People act as if they think you are dishonest.”
- “People act as if they’re better than you are.”
- “You are called names or insulted.”
- “You are threatened or harassed.”

**Options:** a 6-point frequency Options (5 = Almost every day; 4 = At least once a week; 3 = A few times a month; 2 = A few times a year; 1 = Less than once a year; 0 = Never)

**Scoring:** mean item score. The Options is flagged as missing if more than two items do not have a valid score

**Cronbach's alpha (SDoH cohort):** 0.91 [0.904, 0.907]

**Cronbach's alpha (genetics cohort):** 0.91 [0.904, 0.907]

**Code shorthand:** discrimination

#### Stress

**Original source:** Cohen’s perceived stress Options (10-item) [Cohen, Kamarck & Mermelstein 1983](https://www.jstor.org/stable/2136404)

**All of Us source:** [Social Determinants of Health Survey](https://www.researchallofus.org/wp-content/themes/research-hub-wordpress-theme/media/surveys/Survey_SDOH_Eng_Src.pdf)

**Items:**

- “In the last month, how often have you been upset because of something that happened unexpectedly?”
- “In the last month, how often have you felt that you were unable to control the important things in your life?”
- “In the last month, how often have you felt nervous and “stressed”?”
- “In the last month, how often have you felt confident about your ability to handle your personal problems?”
- “In the last month, how often have you felt that things were going your way?”
- “In the last month, how often have you found that you could not cope with all the things that you had to do?”
- “In the last month, how often have you been able to control irritations in your life?”
- “In the last month, how often have you felt that you were on top of things?”
- “In the last month, how often have you been angered because of things that were outside of your control?”
- “In the last month, how often have you felt difficulties were piling up so high that you could

not overcome them?”

**Options:** frequency rating in the past month using a 5-point Options (1 =

Never; 2 = Almost Never; 3 = Sometimes; 4 = Fairly Often; 5 = Very Often)

**Scoring:** multiplying the item mean times ten. It can be scored if more than 8 items have a valid response

**Cronbach's alpha (SDoH cohort):** 0.91 [0.905, 0.908]

**Cronbach's alpha (genetics cohort):** 0.91 [0.905, 0.908]

**Code shorthand:** stress

#### Spiritual Experiences

**Original source:** Fetzer multidimensional measure of religiousness/spirituality (6-item) [Masters, K. S. Brief multidimensional measure of religiousness/spirituality (BMMRS). In Encyclopedia of Behavioral Medicine (edsGellman, M. D. & Turner, J. R.) 267–269 (Springer, 2013)](https://link.springer.com/referenceworkentry/10.1007/978-1-4419-1005-9_1577)

**All of Us source:** [Social Determinants of Health Survey](https://www.researchallofus.org/wp-content/themes/research-hub-wordpress-theme/media/surveys/Survey_SDOH_Eng_Src.pdf)

**Items:**

- “I feel God’s (or a higher power’s) presence”
- “I find strength and comfort in my religion”
- “I feel deep inner peace or harmony”
- “I desire to be closer to or in union with God (or a higher power)”
- “I feel God’s (or a higher power’s) love for me, directly or through others”
- “I I am spiritually touched by the beauty of creation”
- “How often do you go to religious meetings or services?”

**Options:** 6-point Options (6 = Many times a day; 5 = Every day; 4 = Most days; 3 =

Some days; 2 = Once in a while; 1 = Never or almost never) In response to All of Us participant feedback, this response set was altered to add additional choices: 0 = I do not believe in God (or a higher power) or 0 = I am not religious

**Scoring:** average of the items and is flagged as missing if more than two items do not have a valid response

**Cronbach's alpha (SDoH cohort):** 0.83 [0.825, 0.830]

**Cronbach's alpha (genetics cohort):** 0.83 [0.825, 0.830]

**Code shorthand:** spirituality
